# Elevated Liver Damage Biomarkers in Long COVID: A Systematic Review and Meta-Analysis

**DOI:** 10.1101/2024.10.18.24315698

**Authors:** Abbas F. Almulla, Yanin Thipakorn, Yingqian Zhang, Michael Maes

**Author notes:** **Corresponding author** Prof. Dr. Michael Maes, M.D., Ph.D. Sichuan Provincial Center for Mental Health Sichuan Provincial People’s Hospital, School of Medicine, University of Electronic Science and Technology of China Chengdu 610072 China, <u>Michael Maes Google Scholar profile</u>.

## Abstract

**Background:** Long COVID (LC) presents with complex pathophysiology, affecting multiple organs and producing a range of symptoms, from neuropsychiatric disturbances to multi-organ dysfunction. Liver damage has emerged as a notable feature, yet no systematic review or meta-analysis has comprehensively evaluated the biomarkers confirming liver injury in LC patients.

**Objective:** The present study aims to examine blood-based biomarkers of liver damage in LC disease.

**Methods:** A search of PubMed, Google Scholar, SciFinder, and SCOPUS identified 61 eligible studies, including 7172 participants, with 3404 LC patients and 3768 controls.

**Results:** Our analysis identified a significant increase in the liver damage index among LC patients, with a moderate effect size (standardized mean difference, SMD = 0.553; confidence intervals; 95% CI: 0.305–0.760) compared to normal controls. Additionally, LC patients exhibited marked elevations in alanine aminotransferase (SMD = 0.615; 95% CI: 0.351;0.878), aspartate aminotransferase (SMD = 0.352; 95% CI: 0.068;0.637), gamma-glutamyl transferase (SMD = 0.969; 95% CI: 0.194;1.745), and lactate dehydrogenase (SMD = 0.666; 95% CI: 0.332;0.999) activities. Moreover, significant reductions were observed in total protein (SMD = -0.326; 95% CI: -0.631; -0.021) and increases in prothrombin time (SMD = 0.633; 95% CI: 0.077;1.189), ferritin (SMD = 0.437; 95% CI: 0.069;0.805), D-dimer (SMD = 0.359; 95% CI: 0.144;0.573). Further studies are needed to confirm reductions in platelet count and elevations in fibrinogen levels

**Conclusion:** This study suggests that LC is associated with persistent liver damage and coagulopathy, highlighting the need to incorporate liver injury into treatment strategies to reduce potential risks.

## Introduction

The coronavirus disease 2019 (COVID-19), caused by severe acute respiratory syndrome coronavirus 2 (SARS-CoV-2), has posed significant global health and economic challenges since its emergence in late 2019 (World Health Organization 2022). While efforts have primarily focused on reducing acute infection and mortality, a growing body of evidence highlights the persistence of various symptoms long after the resolution of the acute phase (Lopez-Leon, Wegman-Ostrosky et al. 2021, Ewing, Salamon et al. 2024). These lasting symptoms, collectively referred to as Long COVID (LC), can significantly impair the quality of life for many individuals’ months after recovery (Maes, Al-Rubaye et al. 2022). Manifestations of LC range from neuropsychiatric disturbances, such as mood and memory impairments, to systemic multi-organ dysfunction (Fernandez-de-las-Peñas, Notarte et al. 2024, Negrut, Menegas et al. 2024), with the pancreas, heart, liver, kidneys, and lungs being most affected (Dennis, Wamil et al. 2021, Raman, Cassar et al. 2021).

The pathophysiology of LC is complex, involving various biological disturbances, including oxidative and nitrosative stress, immune dysregulation (Al-Hakeim, Al-Rubaye et al. 2023), lowered tryptophan levels, and upregulation of the tryptophan catabolite pathway (Al-Hakeim, Khairi Abed et al. 2023, Almulla, Thipakorn et al. 2024). Other factors include herpes viruses reactivation (Vojdani, Almulla et al. 2024), autoimmune responses (Almulla, Maes et al. 2023, Maes, Almulla et al. 2024), hematological changes such as anemia and lymphopenia, and autonomic dysfunction (Lechuga, Morel et al. 2023, Rinaldi, Rigo et al. 2024). These mechanisms contribute to thrombotic endothelialitis, endothelial inflammation, and fibrinaloid microclots, which affect multiple organ systems (Turner, Khan et al. 2023).

Post-COVID organ damage remains prevalent, with 59-70% of individuals experiencing single-organ impairment and 27-29% having multi-organ impairment 6-12 months after infection (Dennis, Wamil et al. 2021, Dennis, Cuthbertson et al. 2023). Importantly, both symptomatic and asymptomatic individuals may experience organ damage, suggesting that the current definition of LC may require expansion (Ewing, Salamon et al. 2024). Several studies have particularly highlighted liver damage in LC, including hepatocellular injury (Lebbe, Aboulwafa et al. 2024). While hepatitis B and C viruses are the primary contributors, other viruses such as Epstein-Barr virus, cytomegalovirus, and SARS-CoV-2 can also cause liver damage (Adams and Hubscher 2006, Shi, Wang et al. 2023, Ren, Lu et al. 2024).

In COVID-19, elevated liver enzymes—such as alanine transaminase (ALT), aspartate transaminase (AST), and alkaline phosphatase (ALP)—are frequently observed during the acute phase of infection (Ngiam, Chew et al. 2021, Wijarnpreecha, Ungprasert et al. 2021), with approximately 39% of patients showing signs of liver injury (Li, Fan et al. 2023). Severe liver damage in COVID-19 correlates with higher mortality, prolonged hospitalization, and increased morbidity (Nasir et al., 2024). Individuals with pre-existing liver disease may be at greater risk of severe COVID-19 outcomes and further hepatic deterioration (Ali and Hossain 2020, Lee, Kang et al. 2020). Long-term consequences of COVID-19-related liver injury are still under investigation, with potential links to metabolic-fatty liver disease (Nasir et al., 2024). While acute liver injury during COVID-19 is often reversible for many patients, there is emerging evidence that some individuals may experience persistent liver damage even after recovery from the virus. Indeed, emerging evidence suggests these abnormalities may persist or recur in LC (Gameil, Marzouk et al. 2021, Bota, Bratosin et al. 2024), indicating potential ongoing liver dysfunction (Pan, Wang et al. 2024). The mechanisms underlying LC-associated liver injury include direct viral cytotoxicity, cytokine storm, hypoxia, and drug-induced toxicity (Boleslav, Serdyukova et al. 2024, Nasir, Khanum et al. 2024).

We hypothesize that LC shows significant abnormalities in liver damage biomarkers, indicating ongoing damage. No prior meta-analysis has investigated liver enzyme abnormalities and other liver damage indices in LC patients. Therefore, this study aims to assess liver damage biomarkers in LC, including liver damage index, AST/ALT ratio, and individual levels of liver enzymes (AST, ALT, ALP, gamma-glutamyltransferase GGT, lactate dehydrogenase, LDH), albumin, total protein, ferritin, bilirubin, prothrombin time, platelet count, d-dimer, and fibrinogen.

## Materials and method

In this study, we adhered to several key methodological frameworks, including PRISMA 2020 guidelines (Page, McKenzie et al. 2021), the Cochrane Handbook for Systematic Reviews of Interventions, and the Meta-Analyses of Observational Studies in Epidemiology (MOOSE) guidelines (Chandler, Cumpston et al. 2019). Our analysis focused on patients with LC and normal controls, examining liver damage biomarkers such as ALT, AST, GGT, ALP, LDH, albumin, ferritin, bilirubin, as well as prothrombin time, d-dimer, fibrinogen and platelet count. Additionally, we calculated composite indices of liver damage, including the liver damage index and AST/ALT ratio.

### Search strategy

To gather comprehensive data on liver damage biomarkers in long COVID (LC), we conducted a systematic search of electronic databases, including PubMed/MEDLINE, Google Scholar, and SciFinder. This search, covering the period from April 20 to the end of September 2024, was driven by predefined keywords and MeSH terms (see ESF, Table 1). Additionally, we reviewed reference lists from relevant studies and previous meta-analyses to ensure no significant studies were overlooked.

**Table 1.** The outcomes and number of patients with Long COVID (LC) and normal controls along with the side of standardized mean difference (SMD) and the 95% confidence intervals with respect to zero SMD.

| Outcome profiles | n studies | Side of 95% confidence intervals |  |  |  | Patient Cases | Control Cases | Total number of participants |
| --- | --- | --- | --- | --- | --- | --- | --- | --- |
|  |  | < 0 | Overlap 0 and SMD < 0 | Overlap 0 and SMD > 0 | > 0 |  |  |  |
| Liver Damage Index | 30 | 1 | 6 | 15 | 8 | 1890 | 1764 | 3654 |
| AST/ALT ratio | 29 | 3 | 14 | 11 | 1 | 1880 | 1747 | 3626 |
| ALT | 27 | 0 | 6 | 15 | 6 | 1828 | 1653 | 3480 |
| AST | 25 | 1 | 8 | 9 | 7 | 1402 | 1561 | 2962 |
| GGT | 9 | 0 | 3 | 2 | 4 | 413 | 393 | 806 |
| LDH | 25 | 1 | 9 | 5 | 10 | 1718 | 1791 | 3509 |
| ALP | 4 | 0 | 1 | 1 | 2 | 292 | 262 | 554 |
| Albumin | 18 | 5 | 5 | 8 | 0 | 879 | 1184 | 2063 |
| Total protein | 19 | 6 | 5 | 8 | 0 | 938 | 1215 | 2153 |
| Bilirubin | 8 | 0 | 1 | 6 | 1 | 418 | 446 | 864 |
| Ferritin | 28 | 3 | 6 | 12 | 7 | 1707 | 1716 | 3423 |
| Platelet count | 19 | 2 | 9 | 6 | 1 | 1438 | 1429 | 2867 |
| Prothrombin time | 7 | 0 | 0 | 4 | 3 | 321 | 362 | 683 |
| D-dimer | 26 | 0 | 8 | 10 | 8 | 1373 | 1747 | 3119 |
| Fibrinogen | 11 | 0 | 1 | 6 | 4 | 377 | 731 | 1107 |

ALT: Alanine aminotransferase, Aspartate transaminase, ALP: Alkaline phosphatases, GGT: Gamma-glutamyl transferase, LDH: Lactate dehydrogenase. Liver Damage Index: (ALT+ AST+ ALP+GGT+ bilirubin)

### Eligibility criteria

We conducted a comprehensive search for relevant studies, prioritizing peer-reviewed publications in English. However, to maximize data capture, we also included grey literature and studies in Thai, French, Spanish, Turkish, Hungary, German, Ukrainian, Italian, Russian, and Arabic. Studies were included if they met the following criteria: observational design (case-control or cohort) with control groups. Investigation of liver damage biomarkers in LC patients diagnosed based on WHO criteria (World Health Organization 2021). Baseline assessment of biomarkers of liver dysfunction followed by longitudinal follow-up. Exclusion criteria encompassed studies using unconventional biological samples (e.g., hair, saliva, platelet-rich plasma, CSF), focusing on genetic or translational research, or lacking a control group. Furthermore, studies lacking data on mean and standard deviation (SD) or standard error (SE) of biomarkers were excluded unless data could be obtained from the authors upon request or calculated from existing data using established methods and online tools (hkbu.edu.hk, https://automeris.io/WebPlotDigitizer/).

### Primary and secondary outcomes

Our meta-analysis identified several key biomarkers of liver damage, including composite indices like the liver damage index and AST/ALT ratio as shown in Table 1. Beyond these composite measures, we also examined individual biomarkers as secondary outcomes, such as ALT, AST, GGT, bilirubin, LDH, ALP, albumin, total protein, and coagulation factors (prothrombin time, d-dimer, fibrinogen and platelet count).

### Screening and data extraction

The initial screening for this meta-analysis was conducted by two researchers, AA and YT, who evaluated study titles and abstracts according to the predefined inclusion criteria. Subsequent to this initial step, the complete texts of potentially relevant studies were acquired and further assessed, while studies failing to meet the exclusion criteria were eliminated. AA and YT systematically documented essential data in a custom Excel file, including author names, study dates, liver-related biomarker measurements (mean and standard deviation), participant counts in both patient and control groups, and study sample sizes. The file contained supplementary details including study design, sample types (e.g., serum, plasma, cerebrospinal fluid, brain tissue, blood cells), post-COVID timeframes, intensive care unit admissions during the acute illness phase, participant ages, gender distribution, and geographical locations. Discrepancies in data entry or interpretation were resolved through consultations with the senior author, MM.

We assessed the methodological quality of the included studies using the Immunological Confounder Scale (ICS), initially introduced by (Andrés-Rodríguez, Borràs et al. 2020) and subsequently modified by MM for liver damage studies in LC. The ICS comprises two primary instruments: the Quality Scale and the Redpoints Scale, as detailed in Table 4 of the ESF-1. Previous meta-analyses on immune activation and kynurenine pathway in LC (Almulla, Thipakorn et al. 2024, Almulla, Thipakorn et al. 2024), lipid peroxidation and tryptophan metabolism in affective disorders have successfully utilized these scales (Almulla, Thipakorn et al. 2022a, Almulla, Thipakorn et al. 2023). The Quality Scale evaluates factors including sample size, control of confounders, and sampling duration, with scores ranging from 0 (lowest quality) to 10 (highest quality). The Redpoints Scale assesses potential biases in LC immune activation studies, with scores from 0 (optimal control) to 26 (no control).

### Data analysis

The CMA V4 program was used in this meta-analysis in accordance with PRISMA standards (see ESF, Table 2). At least two studies were required for each identified biomarker. We calculated the liver damage index by incorporating relevant biomarkers, including ALT, AST, ALP, GGT, and bilirubin, to account for dependency in the analysis. The AST/ALT was calculated by attributing a positive effect size to AST and a negative effect size to ALT in the comparison of patients to controls, under the assumption of data dependence.

**Table 2.** Results of meta-analysis performed on several outcome variables with combined different media and separately.

| Outcome feature sets | n | Groups | SMD | 95% CI | z | p | Q | df | p | I<sup>2</sup> (%) | $\tau^2$ | T |
| --- | --- | --- | --- | --- | --- | --- | --- | --- | --- | --- | --- | --- |
| Liver Damage Index | 30 | Overall | 0.381 | 0.149;0.613 | 3.214 | 0.001 | 275.284 | 29 | < 0.001 | 89.465 | 0.356 | 0.597 |
| AST/ALT ratio | 29 | Overall | -0.038 | -0.115;0.038 | -0.983 | 0.326 | 29.727 | 28 | 0.376 | 5.809 | 0.003 | 0.050 |
| ALT | 27 | Overall | 0.398 | 0.118;0.677 | 2.784 | 0.005 | 345.228 | 26 | < 0.001 | 92.469 | 0.488 | 0.699 |
| AST | 25 | Overall | 0.352 | 0.068;0.637 | 2.427 | 0.015 | 293.196 | 24 | < 0.001 | 91.814 | 0.466 | 0.682 |
| GGT | 9 | Overall | 0.969 | 0.194; 1.745 | 2.450 | 0.014 | 173.345 | 8 | < 0.001 | 95.385 | 1.317 | 1.148 |
| LDH | 25 | Overall | 0.489 | 0.175;0.802 | 3.055 | 0.002 | 406.721 | 24 | < 0.001 | 94.099 | 0.574 | 0.758 |
| ALP | 4 | Overall | 1.043 | -0.078;2.164 | 1.823 | 0.068 | 96.465 | 3 | < 0.001 | 96.890 | 1.265 | 1.125 |
| Albumin | 18 | Overall | -0.293 | -0.628;0.042 | -1.717 | 0.086 | 179.336 | 17 | < 0.001 | 90.521 | 0.446 | 0.668 |
| Total protein | 19 | Overall | -0.326 | -0.631; -0.021 | -2.095 | 0.036 | 165.345 | 18 | < 0.001 | 89.114 | 0.383 | 0.619 |
| Bilirubin | 8 | Overall | 0.362 | -0.072;0.795 | 1.635 | 0.102 | 60.296 | 7 | < 0.001 | 88.391 | 0.334 | 0.578 |
| Ferritin | 28 | Overall | 0.437 | 0.069;0.805 | 2.328 | 0.020 | 592.304 | 27 | < 0.001 | 95.442 | 0.921 | 0.960 |
| Platelet count | 19 | Overall | -0.226 | -0.584; 0.133 | -1.235 | 0.217 | 321.131 | 18 | < 0.001 | 94.395 | 0.572 | 0.757 |
| Prothrombin time | 7 | Overall | 0.633 | 0.077;1.189 | 2.230 | 0.026 | 63.826 | 6 | < 0.001 | 90.599 | 0.508 | 0.713 |
| D-Dimer | 26 | Overall | 0.359 | 0.144; 0.573 | 3.279 | 0.001 | 182.051 | 25 | < 0.001 | 86.268 | 0.253 | 0.503 |
| Fibrinogen | 11 | Overall | 0.660 | -0.099;1.418 | 1.704 | 0.088 | 273.607 | 10 | < 0.001 | 96.345 | 1.566 | 1.251 |

ALT: Alanine aminotransferase, Aspartate transaminase, ALP: Alkaline phosphatases, GGT: Gamma-glutamyl transferase, LDH: Lactate dehydrogenase. Liver Damage Index: (ALT+ AST+ ALP+GGT+ bilirubin)

A random-effects model utilizing constrained maximum likelihood was employed to aggregate effect sizes. A p-value below 0.05 (two-tailed) was deemed statistically significant, with effect sizes expressed as standardized mean differences (SMD) and 95% confidence intervals (CI). Cohen’s guidelines categorize SMD values of 0.80, 0.50, and 0.20 as large, moderate, and small effects, respectively. Heterogeneity was evaluated through tau-squared statistics, Q, and I², in accordance with prior meta-analyses. Meta-regression was utilized to identify factors contributing to heterogeneity, and subgroup analyses examined biomarker variations across recovery periods and sample types, including serum, plasma, blood cells, and brain tissues.

To determine how stable effect sizes were, sensitivity analyses used the leave-one-out technique. We utilized the fail-safe N technique, continuity-corrected Kendall tau, and Egger’s regression intercept to evaluate publication bias, applying one-tailed p-values for the latter two methods. If Egger’s test indicated asymmetry, the trim-and-fill method was utilized to estimate missing studies and recalculate adjusted effect sizes. Funnel plots were employed to visually evaluate small study effects by mapping study precision against SMD, including both observed and imputed studies.

## Results

### Search Outcomes

In this study, we conducted a systematic search using specific keywords and MeSH terms across databases such as PubMed, Google Scholar, SciFinder, and SCOPUS (see ESF Table 1). As shown in the PRISMA flow chart (**Figure 1**), our initial search retrieved 23572 articles. After a detailed screening to remove irrelevant and duplicate entries, the pool was reduced to 175 studies. Of these, only 61 met the inclusion criteria for our systematic review. Ultimately, 61 studies were included in the meta-analysis, adhering to our specified inclusion and exclusion guidelines (Townsend, Dyer et al. 2020, Aparisi, Ybarra-Falcón et al. 2021, Colarusso, Maglio et al. 2021, Gameil, Marzouk et al. 2021, Kolesova, Vanaga et al. 2021, Taha, Samaan et al. 2021, Wallis, Heiden et al. 2021, Wu, Deng et al. 2021, Alfadda, Rafiullah et al. 2022, Aparisi, Ybarra-Falcón et al. 2022, Belenichev, Kucherenko et al. 2022, Clemente, Sinatti et al. 2022, Corrêa, Deus et al. 2022, Di Gennaro, Valentini et al. 2022, Díaz-Salazar, Navas et al. 2022, Dugani, Mehta et al. 2022, Fan, Wong et al. 2022, Fernández-de-Las-Peñas, Ryan-Murua et al. 2022, García-Abellán, Fernández et al. 2022, Guntur, Nemkov et al. 2022, Kerget, Çelik et al. 2022, Kruger, Vlok et al. 2022, Maamar, Artime et al. 2022, Magdy, Eid et al. 2022, Martone, Tosato et al. 2022, Meisinger, Goßlau et al. 2022, Nádasdi, Sinkovits et al. 2022, Radzina, Putrins et al. 2022, Sasso, Muraki et al. 2022, Sibila, Perea et al. 2022, Stavileci, Özdemir et al. 2022, Sumbalova, Kucharska et al. 2022, Sumbalová, Kucharská et al. 2022, Abdulaziz Alsufyani 2023, Al Masoodi, Radhi et al. 2023, Dudar, Loboda et al. 2023, Garcia-Gasalla, Berman-Riu et al. 2023, Ivchenko, Lobzhanidze et al. 2023, Kalinskaya, Vorobyeva et al. 2023, Kankaya, Yavuz et al. 2023, Kovarik, Bileck et al. 2023, Kuchler, Günthner et al. 2023, Lazebnik, Turkina et al. 2023, Paniskaki, Goretzki et al. 2023, Paris, Palomba et al. 2023, Petramala, Sarlo et al. 2023, Sommen, Havdal et al. 2023, Stufano, Isgrò et al. 2023, Teng, Song et al. 2023, Vollrath, Matits et al. 2023, Yamamoto, Otsuka et al. 2023, Agafonova, Elovikova et al. 2024, Boruga, Septimiu-Radu et al. 2024, Bota, Bratosin et al. 2024, Cezar, Kundura et al. 2024, Frontera, Betensky et al. 2024, Gupta, Nicholas et al. 2024, Parás-Bravo, Fernández-de-las-Peñas et al. 2024, Portacci, Amendolara et al. 2024, Poyatos, Luque et al. 2024, Torki, Hoseininasab et al. 2024).

**Figure 1.**
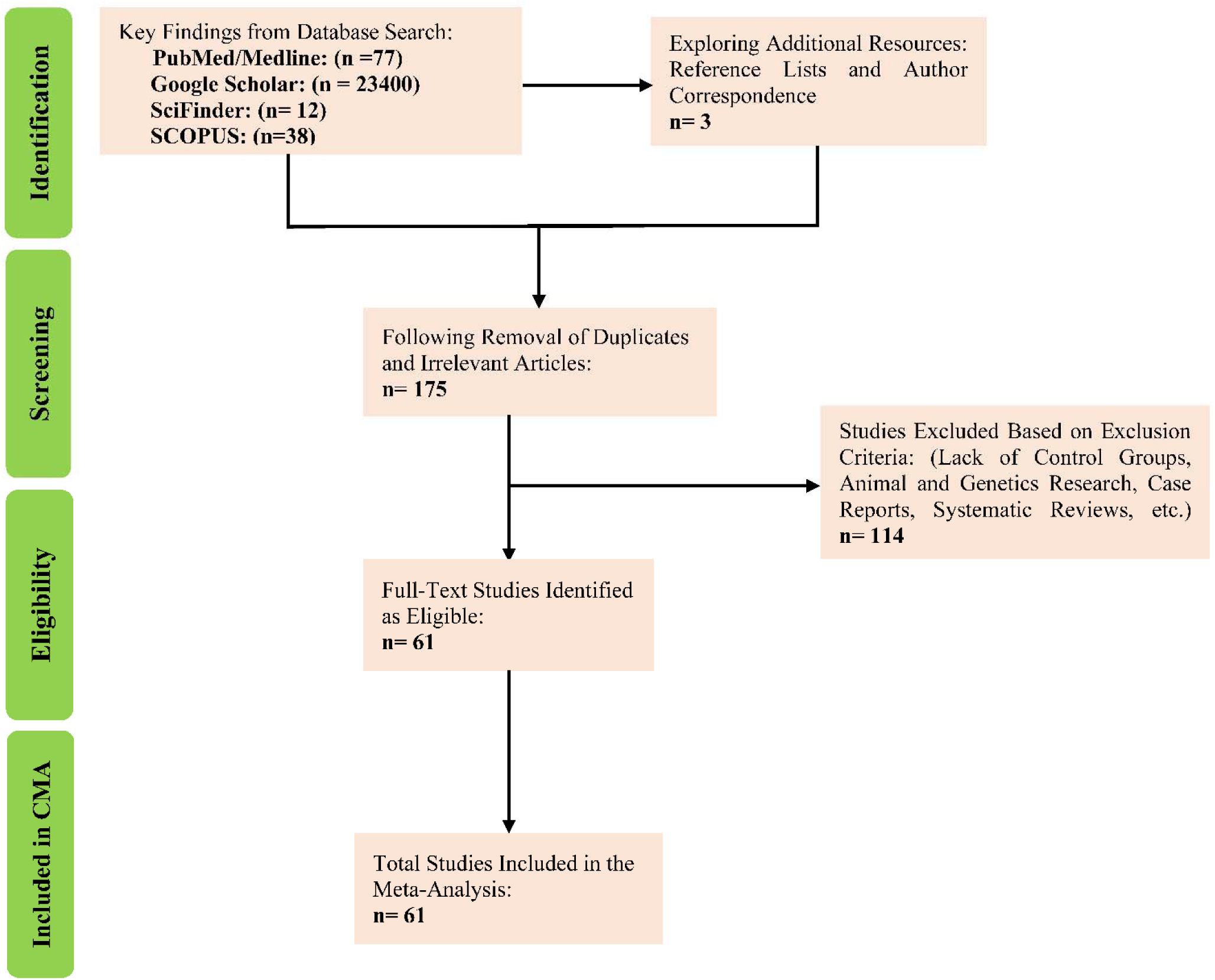
The PRISMA flow chart.

In this meta-analysis, a total of 7172 individuals were included, comprising 3404 LC patients and 3768 control subjects. Participant ages ranged from 11 to 69 years. The majority of studies employed routine laboratory tests in combination with ELISA techniques to assess liver damage biomarkers as detailed in ESF Table 5. A wide range of countries contributed to the dataset, with Spain and Italy leading the contributions with 10 and 8 studies, respectively. Russia and Germany each contributed 4 studies, while Egypt and Turkey each contributed 3. The United States, Ukraine, United Kingdom, Slovakia, Saudi Arabia, Romania, Latvia, and China each provided 2 studies. Additional contributions came from Iraq, Australia, Austria, Brazil, France, Hungary, India, Iran, Ireland, Japan, Norway, Singapore, and South Africa, with 1 study each. The study quality and redpoints were analyzed, with median (min-max) values of 4.5 (min = 2.5, max = 8.5) and 11.5 (min = 4, max = 22.5), respectively. The results are presented in the ESF Table 5.

### Primary outcome variables

#### Liver damage index, and AST/ALT ratio in Long COVID patients versus normal controls

**Table 1** and **Figure 2** indicate that the effect size for the liver damage index computed by 30 studies. Of these, only 1 study reported CIs entirely below zero, while 8 reported CIs entirely above zero. The remaining 21 studies exhibited overlapping CIs, with 6 showing negative SMD values and 15 reporting positive values. As shown in **Table 2**, and **Figure 2**, there is a significant increase in the liver damage index among LC patients compared to NC. The results of publication bias (**Table 3**) revealed 6 studies missing on the right side of the forest plot, and imputing these studies adjusted the SMD value to 0.553.

**Figure 2.**
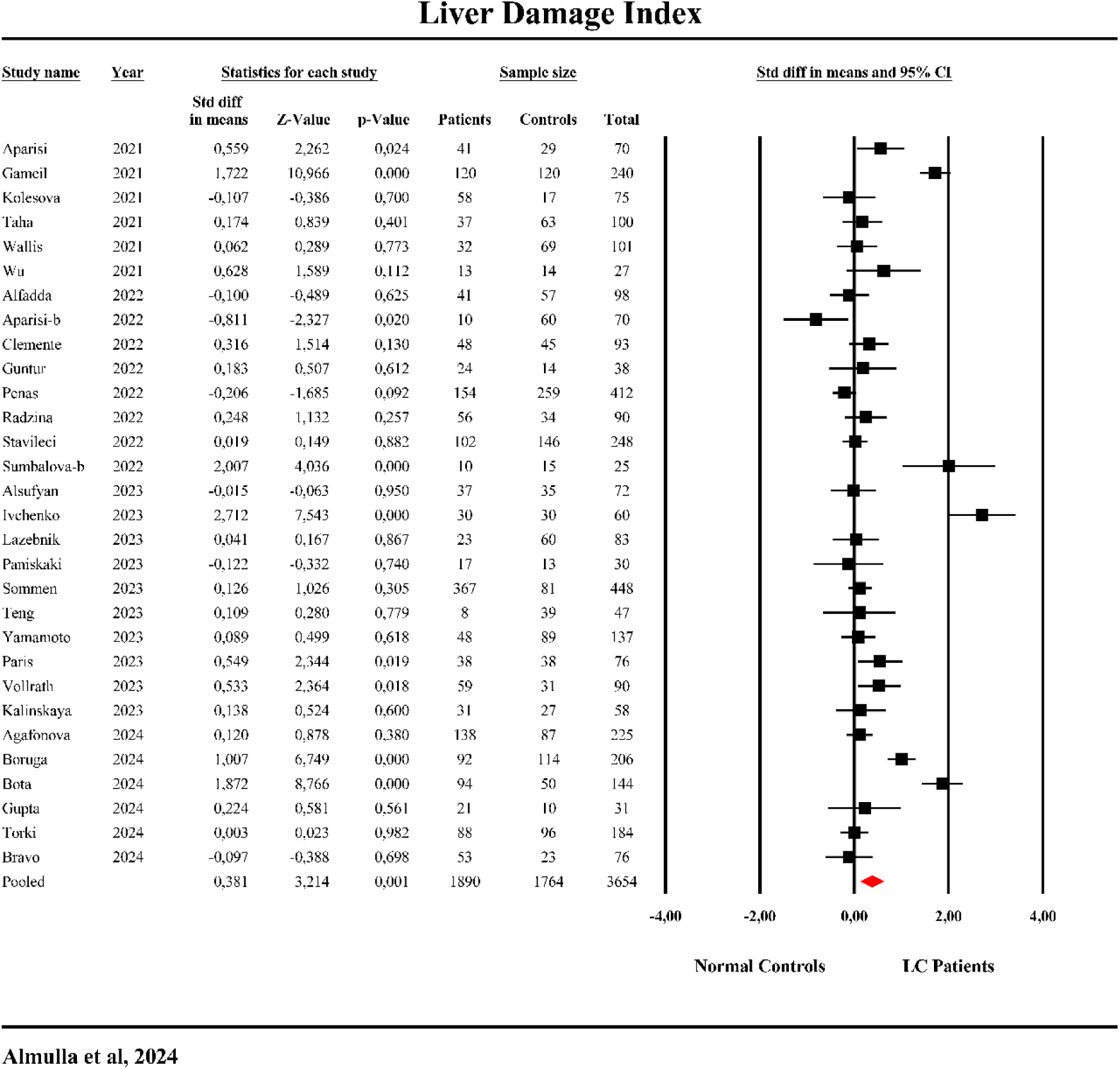
The forest plot of liver damage index in patients with Long COVID (LC) and normal controls.

**Table 3.**
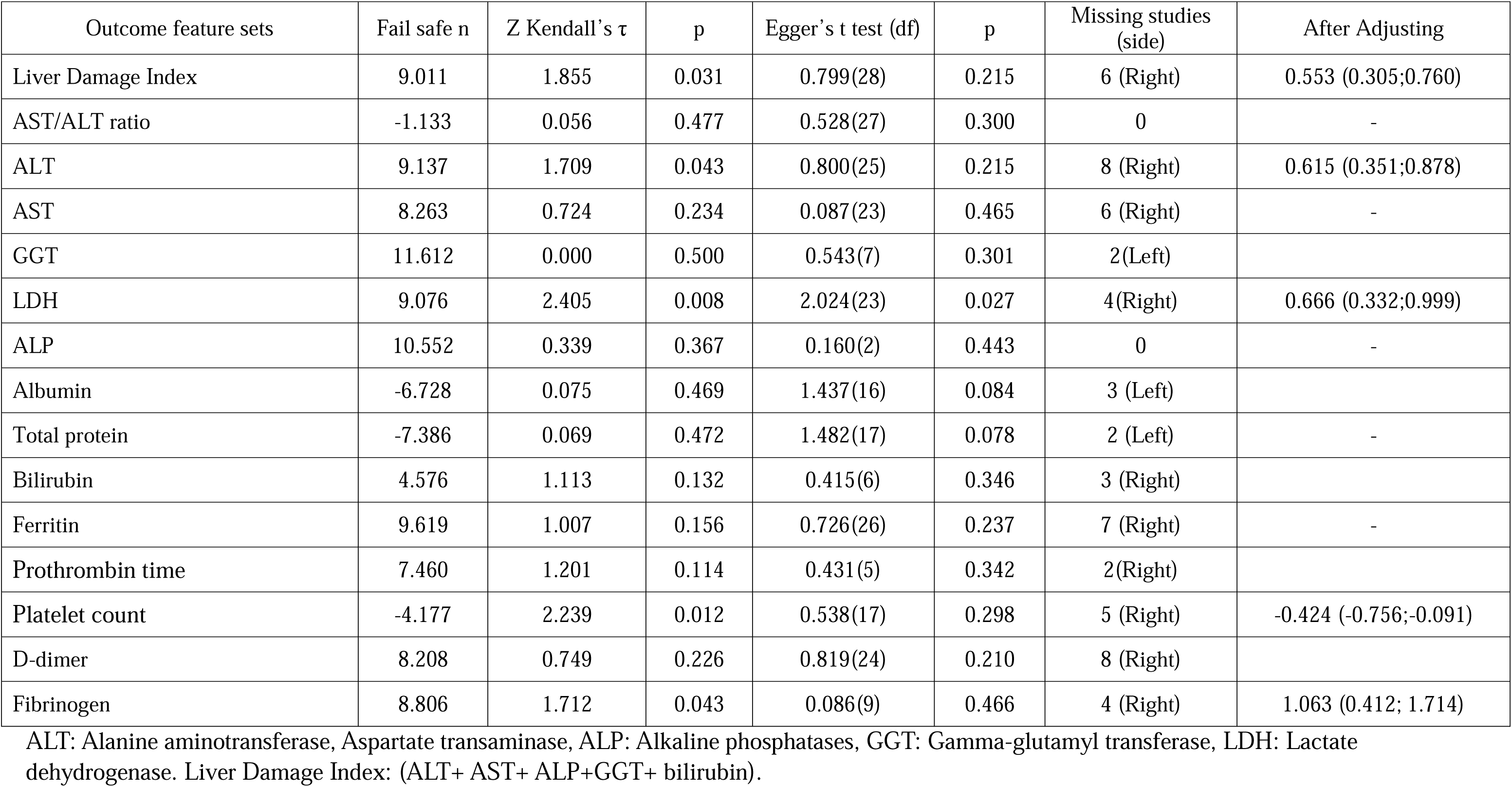
Results on publication bias.

The effect size for the AST/ALT ratio was calculated from 29 studies, as shown in Table 1 and ESF, Figure 1. No significant difference was observed between LC and NC groups (Table 2 and ESF, Figure 1), and Table 3 confirms the absence of bias in these results.

### Secondary Outcome Variables

#### ALT

The effect size for ALT was calculated using data from 27 studies as shown in Table 1 and **Figure 3**. None of the studies reported CIs entirely below zero, while 6 had CIs entirely above zero. The remaining 21 studies showed overlapping CIs, with 6 reporting negative SMD values and 15 showing positive values. Both Figure 3 and Table 2 demonstrate a significant increase in ALT enzyme activity in LC patients compared to NC. As shown in Table 3, publication bias analysis identified 8 missing studies on the right side of the funnel plot; imputing these studies raised the SMD value to 0.615.

**Figure 3.**
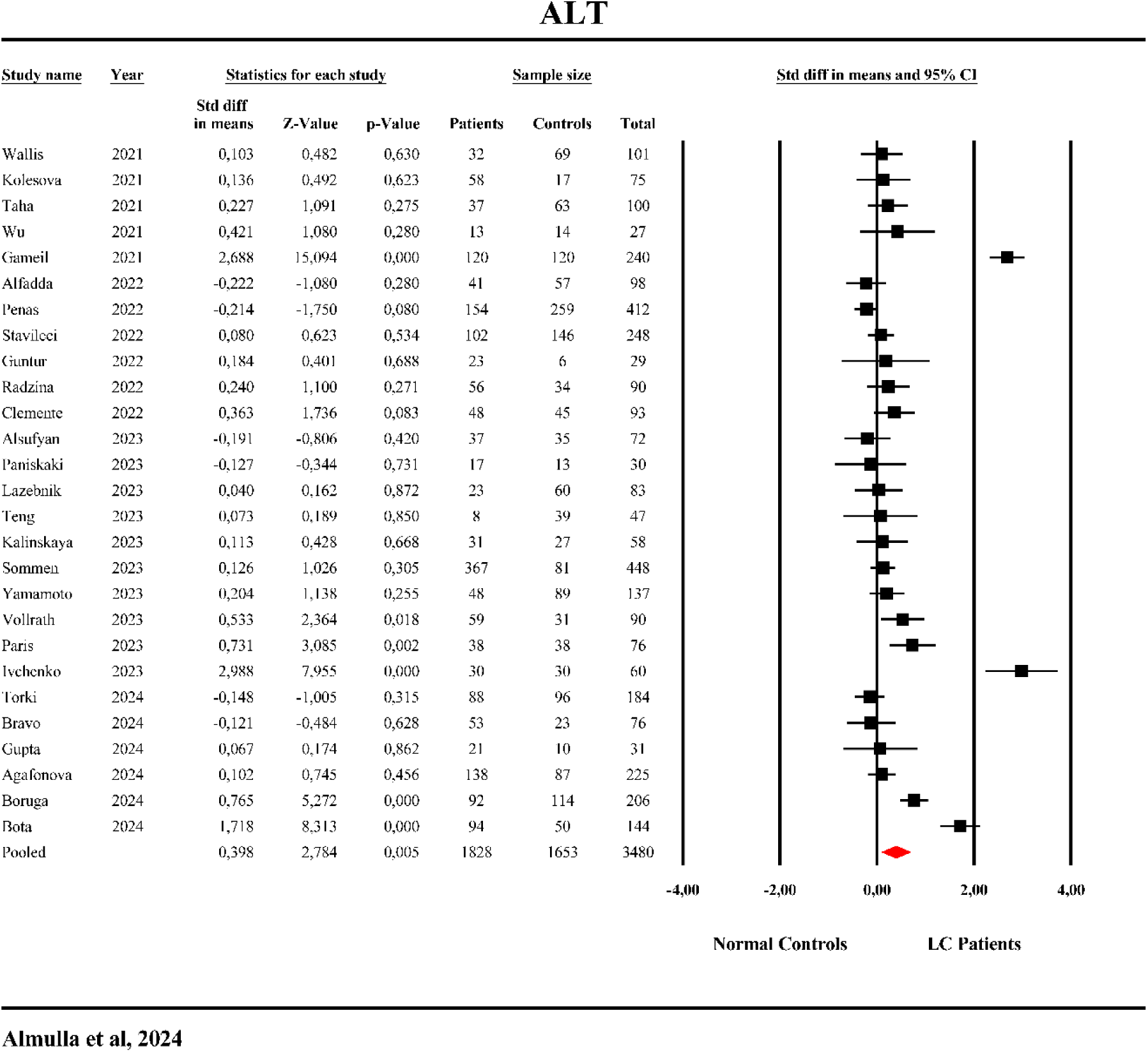
The forest plot of alanine aminotransferase (ALT) in patients with Long COVID (LC) and normal controls.

#### AST

The AST effect size was calculated from 25 studies as displayed in Table 1 and **Figure 4**. Of these, only one study had CIs entirely below zero, while 7 had CIs entirely above zero. Seventeen studies showed overlapping CIs, with mixed results: 8 reported negative SMD values, and 9 had positive SMD values. As shown in Table 2 and Figure 4, AST levels were significantly elevated in LC patients with a moderate SMD compared to NC. Bias tests (Kendall’s and Egger’s) indicated no significant bias in these results.

**Figure 4.**
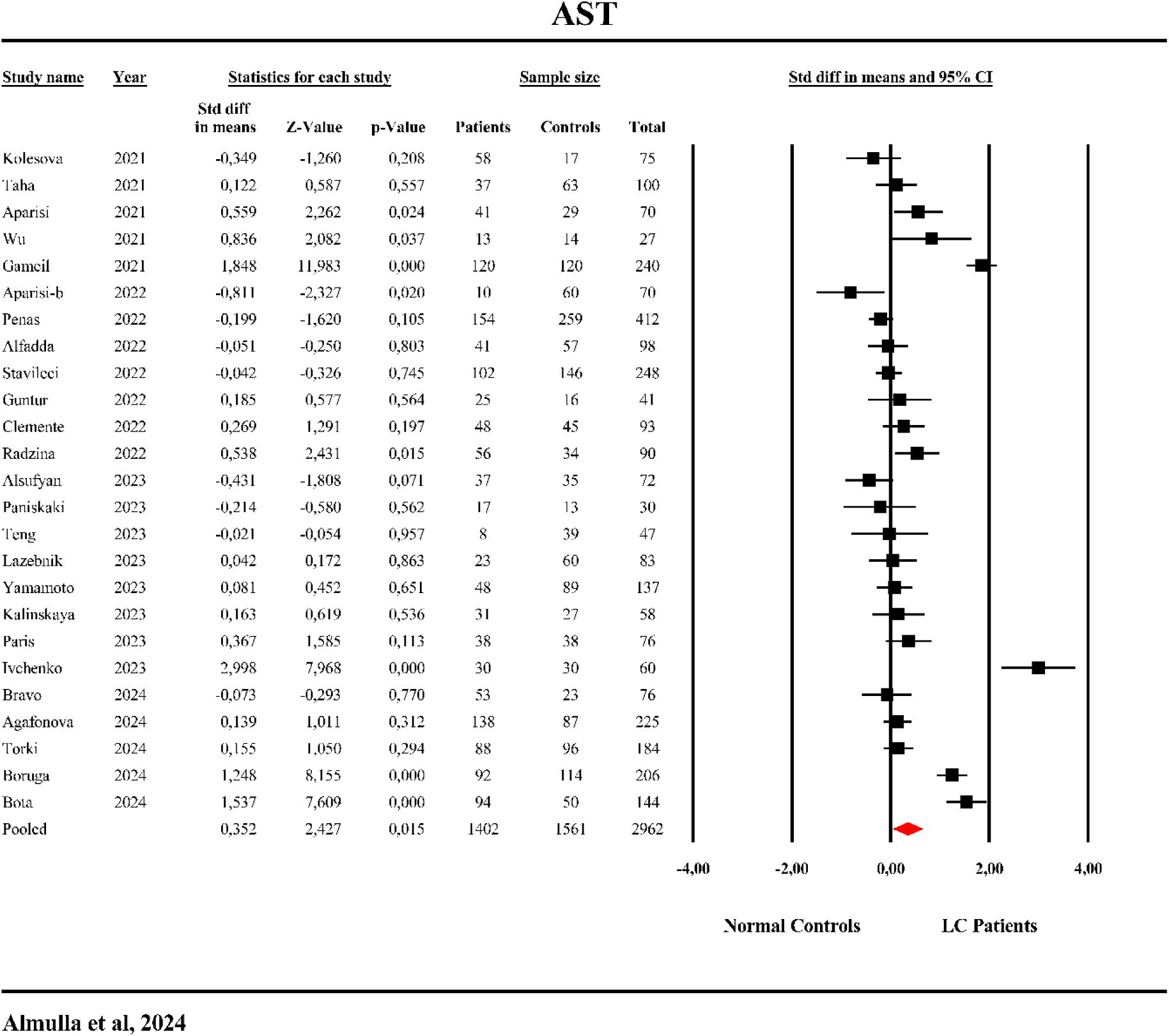
The forest plot of aspartate aminotransferase (AST) in patients with Long COVID (LC) and normal controls.

#### ALP

The effect size of ALP was based on 4 studies (see Table 1 and ESF, Figure 2). No significant differences were observed between LC and NC patients. Table 3 showed no bias.

#### GGT

GGT effect size was calculated from 9 studies as shown in Table 1 and **Figure 5**. None of the studies reported CIs entirely below zero, while 4 had CIs entirely above zero. Five studies demonstrated overlapping CIs, with 3 reporting negative SMD values and 2 positive values. Table 2 and Figure 5 show that GGT levels were significantly elevated in LC patients compared to NC. **Table 3** revealed no bias.

**Figure 5.**
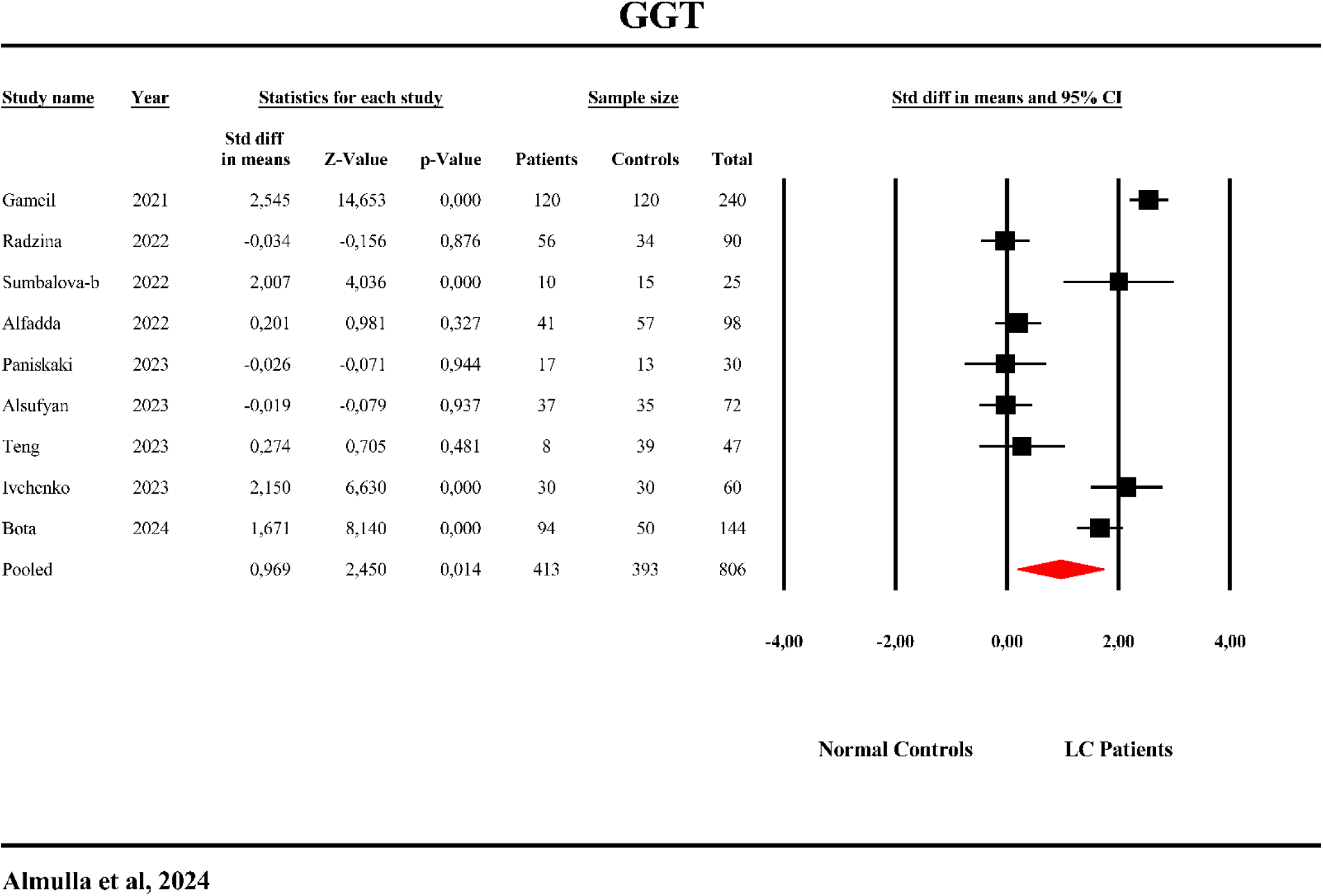
The forest plot of gamma-glutamyl transferase (GGT) in patients with Long COVID (LC) and normal controls.

#### LDH

Table 1 and ESF, Figure 3 indicate that the LDH effect size was computed from 25 studies. Only 1 study reported CIs entirely below zero, while 10 had CIs entirely above zero. The remaining 14 studies showed overlapping CIs, with nine reporting negative SMD values and five positive values. Figure 5 and Table 2 demonstrate a significant increase in LDH activity in LC patients compared to NC. No significant bias was observed (**Table 3**).

#### Albumin

Albumin effect size was calculated from 18 studies, as demonstrated in Table 1 and ESF, Figure 4. No significant differences in albumin levels were found between LC and NC patients (Table 2). Table 3 indicates that publication bias was not detected in the albumin results.

#### Total Protein

The effect size for total protein was derived from 19 studies (see Table 1 and **Figure 6**). Of these, 6 studies reported CIs fully below zero, while none reported CIs fully above zero. Thirteen studies presented overlapping CIs, with 5 indicating negative SMD values and 8 showing positive values. As shown in Table 2 and Figure 6, total protein levels were significantly lower in patients with LC compared to NC. An analysis of publication bias (Table 3) revealed no significant indication of bias.

**Figure 6.**
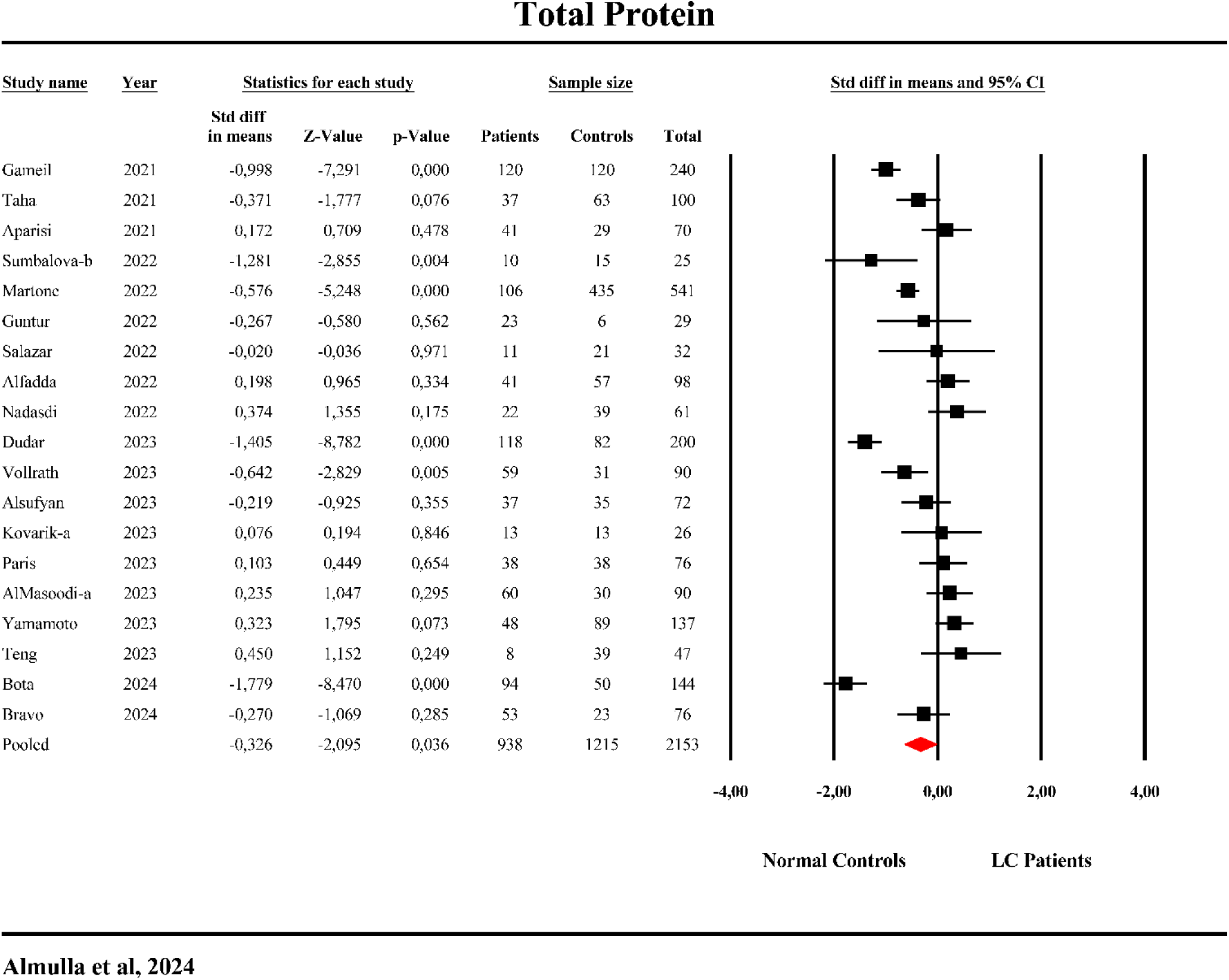
The forest plot of total protein in patients with Long COVID (LC) and normal controls.

#### Bilirubin

Bilirubin effect size was estimated from 8 studies, as shown in Table 1 and ESF, Figure 5. Of these studies, none had CIs entirely below zero, and 1 had CIs entirely above zero. The remaining 7 studies displayed overlapping CIs, with 1 showing negative SMD values and 6 reporting positive ones. Table 2 and ESF Figure 5 reveal that bilirubin levels were significantly lower in LC patients compared to NC. Table 3 showed no substantial evidence of publication bias.

#### Ferritin

Ferritin effect size was pooled from 28 studies (Table 1 and ESF, Figure 6). Among these, 3 studies reported CIs entirely below zero, while 7 had CIs entirely above zero. Eighteen studies exhibited overlapping CIs, with 6 showing negative SMD values and 12 reporting positive values. Ferritin levels were significantly higher in LC patients compared to NC, as shown in Table 2 and Figure 6. No significant bias was found in these results.

#### Platelet count

The effect size for platelet count was derived from 19 studies, as indicated in Table 1 and ESF, Figure 7. Two studies reported CIs entirely below zero, while one study had CIs entirely above zero. Fifteen studies showed overlapping CIs, with 9 reporting negative SMD values and 6 positive values. No significant difference in platelet count was observed between LC and NC patients, as shown in Table 2 and ESF, Figure 7. However, the results of bias analysis in Table 3 identified 5 missing studies on the right side of the funnel plot, and imputing these studies shifted the results toward a significantly decreased platelet count.

**Figure 7.**
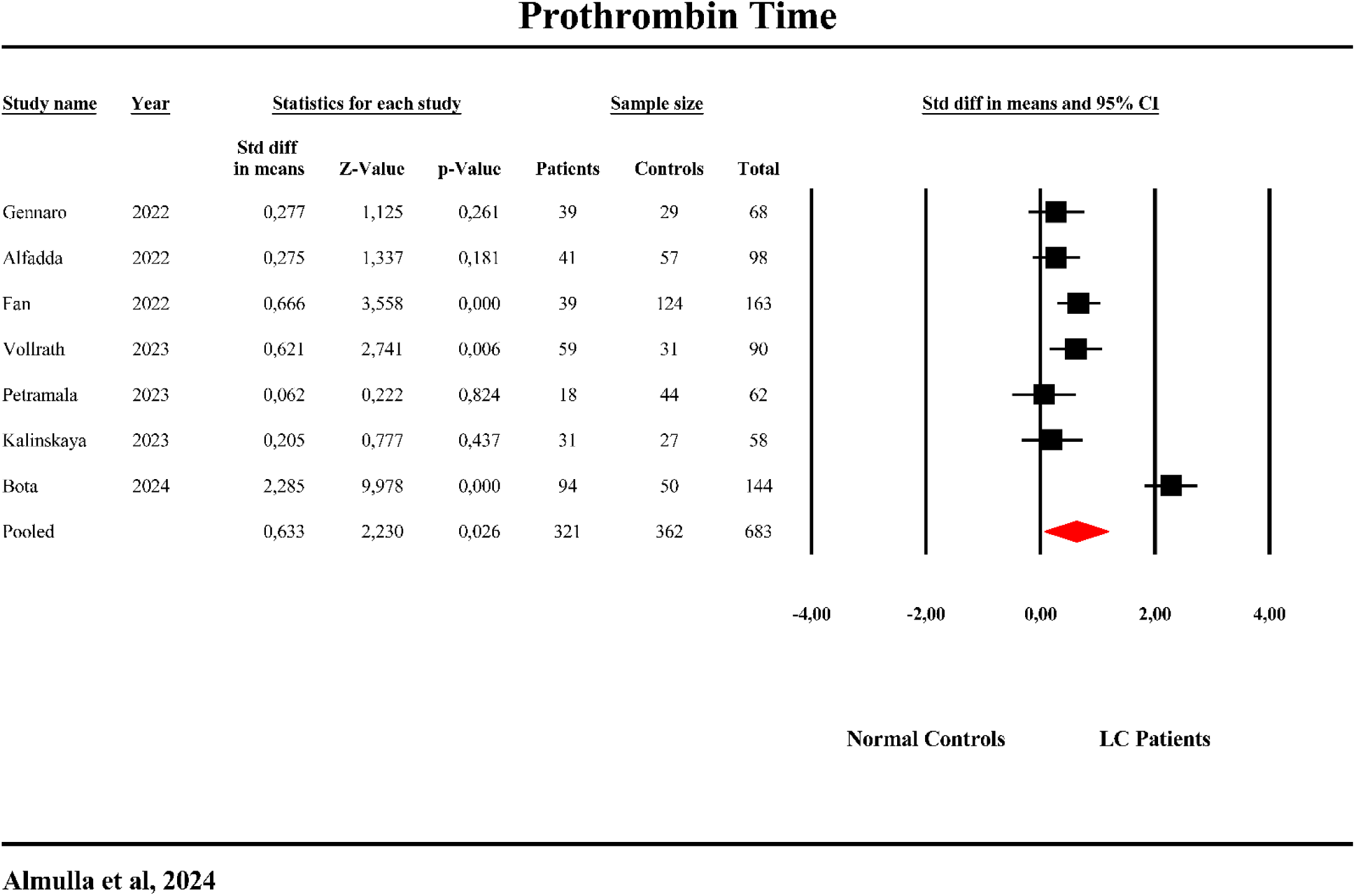
The forest plot of total protein in patients with Long COVID (LC) and normal controls.

#### Prothrombin time

The effect size for prothrombin time was calculated from 7 studies (see Table 1 and **Figure 7**). No studies reported CIs entirely below zero, while 3 study had CIs entirely above zero. Four studies showed overlapping CIs, with 0 reporting negative SMD values and 4 positive values. Table 2 and Figure 8 show that prothrombin time was significantly increased in LC patients compared to NC. No significant differences were observed in prothrombin time between LC and NC patients, as shown in Table 2 and ESF, Figure 6, and no publication bias was detected (see Table 3).

**Figure 8.**
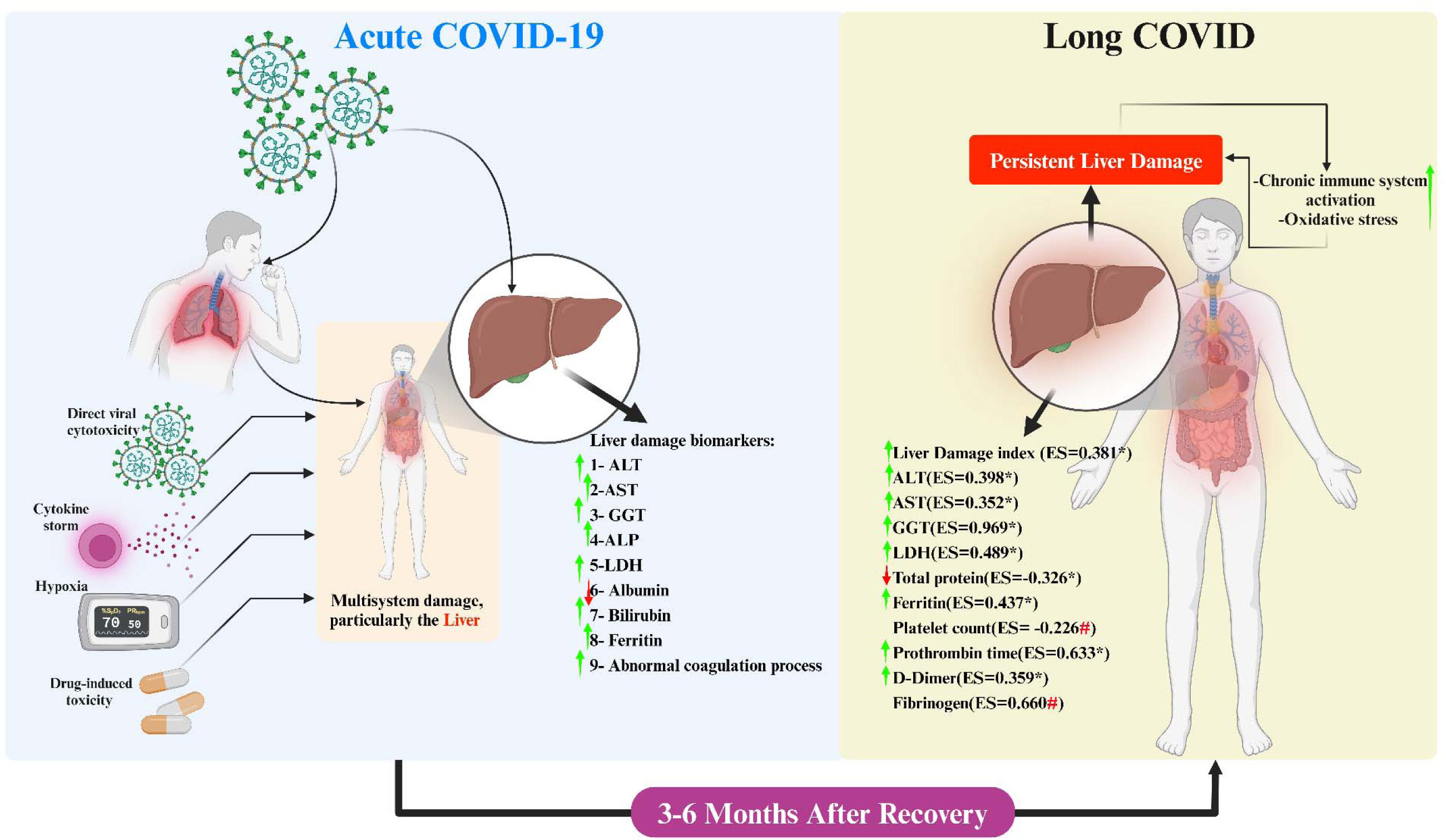
Summary of the liver damage in Long COVID (LC) disease. ALT: Alanine aminotransferase, Aspartate transaminase, ALP: Alkaline phosphatases, GGT: Gamma-glutamyl transferase, LDH: Lactate dehydrogenase, ES: Effect size. *: Indicate a significant difference, #: Publication bias indicate missing studies.

#### D-dimer

D-dimer effect size was derived from a total of 26 studies, as shown in Table 1 and ESF, Figure 8. Notably, none of the studies had CIs completely below zero, while 8 studies had CIs entirely above zero. The remaining 18 studies exhibited overlapping CIs, with 8 showing negative SMD values and 10 displaying positive values. Table 2 and ESF Figure 7, show that D-dimer levels were significantly lower in patients with LC compared to NC. Table 3 indicated no significant bias in the results.

#### Fibrinogen

Table 1 and ESF Figure 9 show that fibrinogen effect size was calculated from 11 studies. None of these studies had CIs fully below zero, while 4 studies had CIs entirely above zero. The remaining 7 studies demonstrated overlapping CIs, with 1 reporting a negative SMD and 6 showing positive values. Fibrinogen levels did not exhibit a significant difference between LC patients and NC as shown in Table 2 and ESF Figure 9. Nevertheless, Kendall and Egger tests in Table 3 revealed the absence of four studies on the right side of the funnel plot. After imputing these missing studies, the results shifted, indicating a significant increase in fibrinogen levels.

### Meta-regression analysis

In this study, we performed a meta-regression analysis (ESF, Table 6) to detect the factors contributing to the heterogeneity observed in studies investigating liver dysfunction biomarkers in LC patients. The analysis revealed that smoking influenced most of the outcomes including ALT, LDH, albumin, total protein and d-dimer. Moreover, using of immunomodulatory drugs shows significant impact on AST/ALT ratio, albumin, and total protein. Additionally, hospitalization during the acute COVID stage significantly influenced several outcomes, including LDH, platelet count, and D-dimer. Post-COVID period had a significant impact on the outcomes for example less than 3 months influenced ferritin levels and platelet count while more than 6 months affected the fibrinogen level. Other variables, such as age, BMI, sample size, quality control, and red points scores, were also found to significantly affect some of the outcomes, as detailed in ESF, Table 6.

## Discussion

This study is the first to conduct a meta-analysis and meta-regression exploring liver damage in LC patients, based on blood-based biomarkers such as liver enzymes, proteins, and indicators of coagulation issues.

### Elevated Liver Indices and Enzyme Activities Indicative of Liver Injury in Long COVID

One of the main findings of this study is that LC patients show significantly higher liver damage index and AST/ALT ratio compared to normal controls. In addition, enzymes like AST, ALT, GGT, and LDH were elevated, suggesting that liver injury can persist for 3–6 months after the initial COVID-19 infection. The ALP enzyme levels did not show significant differences between the two groups. These findings align with those of Gameil et al. (2021) and Bota et al. (2024), who reported ongoing liver enzyme elevations, raising concerns about long-term liver damage, including fibrosis (Gameil, Marzouk et al. 2021, Bota, Bratosin et al. 2024). Ivchenko et al. (2023) also highlighted a link between elevated liver enzymes and cognitive issues, indicating that liver dysfunction in LC may have wide-ranging effects on both physical and mental health (Ivchenko, Lobzhanidze et al. 2023). A recent meta-analysis detected high levels of LDH in post COVID patients compare to normal convalescents (Yong, Halim et al. 2023).

The persistence of elevated liver enzymes might point to ongoing inflammation or early-stage fibrosis, even when patients appear asymptomatic (Malakouti, Kataria et al. 2017). Previous research, suggests that long-term liver complications could go unnoticed until more serious issues arise (Hossain, Ahamed et al. 2024), making regular monitoring crucial for LC patients. In particular, GGT elevations, often associated with oxidative stress (Lim, Yang et al. 2004), may play a significant role in the prolonged liver damage seen in these patients. Al-Hakeim et al. reported that LC is characterized by high oxidative and nitrosative stress as indicated by high levels of their products including malonaldehyde, nitric oxide, myeloperoxidase enzyme (Al-Hakeim, Al-Rubaye et al. 2023). Elevated ALT levels, rather than AST, have been specifically associated with insulin resistance, as demonstrated in prior studies (Simental-Mendía, Rodríguez-Morán et al. 2017, Yamamoto, Prado-Núñez et al. 2020), and insulin resistance itself is a recognized risk factor for depression. In a recent study on LC, we identified a significant increase in insulin resistance, which emerged as a strong predictor of depressive symptoms in these patients (Al-Hakeim, Al-Rubaye et al. 2023).

Elevated ALT, commonly a marker for NAFLD, was found to be a significant independent predictor of both minor and major depression. This finding suggests that ALT elevation may reflect underlying hepatic inflammation associated with NAFLD, which could play a role in the development of depressive symptoms (Zelber-Sagi, Toker et al. 2013). Furthermore, even in healthy adult individuals, higher GGT are connected to negative mood states (Savage, Porter et al. 2023). This suggests that liver injury may contribute to the fatigue and mood disturbances seen in LC patients. Liver-brain interactions during inflammatory liver diseases influence mood through the release of cytokines like TNF-α, IL-1β, and IL-6, which activate central immune responses and neuroinflammation (D’Mello and Swain 2014). This affects neurotransmitter systems, particularly serotonergic and corticotropin-releasing hormone pathways, leading to mood disturbances such as depression and anxiety (D’Mello and Swain 2014) reinforcing the idea that LC is a multi-organ condition.

In patients who experienced severe COVID-19, AST levels were shown to correlate with higher mortality, disease severity, and longer hospital admission (Boregowda, Aloysius et al. 2020, Gu, Li et al. 2020). Our meta-regression analysis showed a negative association between the number of hospitalized patients during the acute COVID-19 stage and LDH levels, platelet counts, and D-dimer. Moreover, these severe patients often had elevated markers of systemic inflammation, such as IL-6, CRP, ferritin, and LDH (Effenberger, Grander et al. 2021), which persisted beyond the acute infection phase (Dugani, Mehta et al. 2022, Almulla, Thipakorn et al. 2024). This prolonged inflammation could explain why liver dysfunction continues long after recovery, as seen in this study. Our meta-regression revealed a strong association between smoking and elevated liver injury biomarkers (ALT, LDH, and D-dimer), highlighting its negative impact on liver health. This is consistent with previous studies on smoking’s harmful effects on liver function (Wannamethee and Shaper 2010).

The mechanisms behind liver injury in LC are complex. SARS-CoV-2 can infect liver cells directly through angiotensin-converting enzyme 2 (ACE2) and TMPRSS2 receptors on these cells, triggering an immune response that leads to inflammation and liver damage (Lyalyukova, Dolgalev et al. 2021, Lebbe, Aboulwafa et al. 2024). Additionally, COVID-19 treatments, such as antivirals and antibiotics, can have toxic effects on the liver, contributing to the damage (Papadopoulos, Vasileiadi et al. 2020). Mitochondrial dysfunction in liver cells may also worsen this inflammation (Akbari and Taghizadeh-Hesary 2023). Those with pre-existing liver conditions like NAFLD face an even higher risk of severe liver damage post-COVID (Hu, Sun et al. 2022), making it essential to monitor these patients closely to prevent complications like liver fibrosis (Caballero-Alvarado, Zavaleta Corvera et al. 2023).

### Hypoproteinemia, and Hyperferritinemia in Long COVID

Another key finding from this study is that LC patients tend to have lower levels of serum total protein and elevated ferritin levels. This is another evidence suggesting ongoing liver dysfunction in LC that reduces the liver’s ability to produce circulating proteins. However, albumin did not show significant change in LC patients compared to normal controls which is probably due to the long half-life of albumin which requires more time to be affected which can appear in advanced stage of liver injury. Studies by Gameil et al. (2021) and Dudar et al. (2023) show similar results (Gameil, Marzouk et al. 2021, Dudar, Loboda et al. 2023). Pasini et al. (2021) also noted that these changes might point to altered metabolism and a hypercatabolic state, where the body breaks down more tissue than it builds (Pasini, Corsetti et al. 2021).

Ferritin could serve as a marker for predicting fatigue and cognitive issues, particularly brain fog, which is common in LC patients (Ishikura, Nakano et al. 2023, Yamamoto, Otsuka et al. 2023). Elevated ferritin levels in major depression are linked to the inflammatory processes and aberrations in iron metabolism in depression (Maes, Van de Vyvere et al. 1996). This means that the fatigue and depressive symptoms in LC could partly stem from the liver’s inability to regulate protein and iron metabolism properly (Huang and Lee 2007). In LC disease, elevated IL-6 (Almulla, Thipakorn et al. 2024), which stimulates hepcidin production, can reduce iron export and increase ferritin levels, creating a state of iron overload (Yilmaz, Eren et al. 2021). These changes in iron metabolism might also contribute to the fatigue and other persistent symptoms observed in LC(Portugal-Nunes, Castanho et al. 2020).

Low albumin levels, commonly seen in COVID-19, are linked to higher mortality rates and increased inflammation (Aziz, Fatima et al. 2020). Additionally, reduced albumin has been associated with depressive and anxiety symptoms, likely due to broader nutritional and inflammatory responses (Maes, Vandewoude et al. 1991), which may be triggered by SARS-CoV-2 infection or its consequences(Al-Jassas, Al-Hakeim et al. 2022). This study’s findings, therefore, suggest that LC patients may suffer from a state of chronic inflammation, which could affect both mental and physical health. Decreased albumin and total protein levels in LC may also be influenced by kidney impairment (Boruga, Septimiu-Radu et al. 2024), hormonal imbalances (Sunada, Honda et al. 2022), and secondary infections, like Clostridioides difficile (Korsunskiy, Belousova et al. 2023).

### Coagulation Abnormalities and Their Implications in Long COVID-Related Liver Injury

A significant finding of this study is that LC patients show elevated levels of D-dimer and no significant changes in fibrinogen and platelet counts. However, publication bias analysis suggests that additional studies are needed to confirm potential increases in fibrinogen and decreases in platelet counts. These results suggest ongoing coagulation abnormalities due to liver injury in LC disease. Interestingly, this study found significant increases in PT in LC patients indicating that those patients are prone to bleeding and hemorrhage. These findings are consistent with other studies that have shown similar disturbances in LC patients (Gameil, Marzouk et al. 2021, Dugani, Mehta et al. 2022, Fan, Wong et al. 2022). Elevated D-dimer levels suggest that blood clotting issues persist well beyond the acute phase of the infection, potentially increasing the risk of microvascular clots and organ damage, including the liver (Levi, Thachil et al. 2020). In LC disease, higher D-dimer levels are linked with more severe and prolonged symptoms, including fatigue, cognitive impairment, and respiratory issues (Acanfora, Nolano et al. 2022, Nicolai, Kaiser et al. 2023).

The presence of microclots, which are resistant to being broken down, has been documented in LC patients (Walker, Federico et al. 2022). These clots can block small blood vessels, contributing to symptoms like fatigue and cognitive dysfunction (Pretorius, Vlok et al. 2021). Increased levels of hemolysis markers and complement system dysfunction further support the role of thromboinflammation in LC pathology (Cervia-Hasler, Brüningk et al. 2024). These ongoing coagulation disturbances may contribute to multi-organ issues, especially in the cardiovascular system (Acanfora, Nolano et al. 2022). Elevated D-dimer levels may also serve as markers for predicting the severity of LC symptoms, including the risk of thrombotic events (Nicolai, Kaiser et al. 2023).

The persistent coagulation abnormalities observed highlight the intricate relationship between liver function, inflammation, and vascular damage (Talla, Vasaikar et al. 2022). Microvascular thrombosis, which is worsened by pre-existing conditions, may reduce blood flow to the liver, leading to ischemic injury (Marín-Gómez, Bernal-Bellido et al. 2012). Viral infections can disrupt the entire blood clotting process, potentially causing both bleeding and clotting issues (Goeijenbier, van Wissen et al. 2012). LC is characterized by a state of increased blood clotting, which raises the risk of stroke and other serious conditions (Kruger, Vlok et al. 2022). Given these risks, therapies that focus on managing clotting dysfunction, such as anticoagulants or anti-inflammatory treatments, may be beneficial in restoring balance and preventing further complications (Kruger, Vlok et al. 2022).

## Limitations

Several limitations must be considered when interpreting our findings. First, the liver is highly vulnerable to damage from various medications, but most of the studies included in this meta-analysis did not provide sufficient data on medication use whether in acute or post-acute COVID stage. This limits our ability to assess how medications may have influenced the results. Additionally, the findings would be more comprehensive if we could have included oxidative and nitrosative stress biomarkers, given their strong association with liver enzymes in LC. Furthermore, since transaminase levels can be elevated due to conditions beyond liver damage, future research should monitor these enzymes while accounting for comorbidities such as cardiovascular, muscle, and thyroid disorders. Lastly, we were unable to evaluate the impact of alcohol consumption on liver enzyme levels, as the studies did not provide adequate information on alcohol use in LC patients, despite its known effect on liver health (Nivukoski, Bloigu et al. 2019).

## Conclusion

**Figure 8** summarizes this study’s findings, suggesting that LC is associated with ongoing liver damage and coagulopathy. The findings highlight that LC involves multiple organ systems, which may lead to serious complications if not closely monitored. Future treatment strategies should take liver damage into account to prevent potential adverse outcomes.

## Data Availability

The corresponding author (MM) will address any reasonable requests for access to the dataset (Excel file) used in this meta-analysis,once all authors have fully utilized the data.

## Ethical approval and consent to participate

Not applicable.

## Consent for publication

Not applicable.

## Availability of data and materials

The corresponding author (MM) will address any reasonable requests for access to the dataset (Excel file) used in this meta-analysis, once all authors have fully utilized the data.

## Author’s contributions

AA and MM developed the study design, with AA, YT, and YZ working together to collect the data. AA and MM carried out the statistical analysis. All authors collaborated in writing the paper and gave their full approval for submitting the final version.

## Acknowledgments

Not applicable.

## References

Abdulaziz Alsufyani, A. (2023). "Post-COVID-19 effect on biochemical parameters in children: Should we take heed?" Saudi J Biol Sci 30(5): 103649.

Acanfora, D., M. Nolano, C. Acanfora, C. Colella, V. Provitera, G. Caporaso, G. R. Rodolico, A. S. Bortone, G. Galasso and G. Casucci (2022). "Impaired Vagal Activity in Long-COVID-19 Patients." Viruses 14(5).

Adams, D. H. and S. G. Hubscher (2006). "Systemic viral infections and collateral damage in the liver." Am J Pathol 168(4): 1057–1059.

Agafonova, T., N. Elovikova, O. Bronnikova and D. Golyadinets (2024). "Post-COVID Syndrome: Persistence of Symptoms and Risk Factors (Longitudinal Observational Study)." The Russian Archives of Internal Medicine 14: 108–115.

Akbari, H. and F. Taghizadeh-Hesary (2023). "COVID-19 induced liver injury from a new perspective: Mitochondria." Mitochondrion 70: 103–110.

Al-Hakeim, H. K., H. T. Al-Rubaye, D. S. Al-Hadrawi, A. F. Almulla and M. Maes (2023). "Long-COVID post-viral chronic fatigue and affective symptoms are associated with oxidative damage, lowered antioxidant defenses and inflammation: a proof of concept and mechanism study." Molecular Psychiatry 28(2): 564–578.

Al-Hakeim, H. K., H. T. Al-Rubaye, A. S. Jubran, A. F. Almulla, S. R. Moustafa and M. Maes (2023). "Increased insulin resistance due to Long COVID is associated with depressive symptoms and partly predicted by the inflammatory response during acute infection." Braz J Psychiatry.

Al-Hakeim, H. K., A. Khairi Abed, S. Rouf Moustafa, A. F. Almulla and M. Maes (2023). "Tryptophan catabolites, inflammation, and insulin resistance as determinants of chronic fatigue syndrome and affective symptoms in long COVID." Frontiers in Molecular Neuroscience 16.

Al-Jassas, H. K., H. K. Al-Hakeim and M. Maes (2022). "Intersections between pneumonia, lowered oxygen saturation percentage and immune activation mediate depression, anxiety, and chronic fatigue syndrome-like symptoms due to COVID-19: A nomothetic network approach." J Affect Disord 297: 233–245.

Al Masoodi, W. T. M., S. W. Radhi, H. K. Abdalsada and H. K. Al-Hakeim (2023). "Correlation between Galanin and its receptor with the serum electrolytes in Long-COVID patients." medRxiv: 2023.2011.2027.23299076.

Alfadda, A. A., M. Rafiullah, M. Alkhowaiter, N. Alotaibi, M. Alzahrani, K. Binkhamis, K. Siddiqui, A. Youssef, H. Altalhi, I. Almaghlouth, M. Alarifi, S. Albanyan, M. F. Alosaimi, A. Isnani, S. S. Nawaz and K. Alayed (2022). "Clinical and biochemical characteristics of people experiencing post-coronavirus disease 2019-related symptoms: A prospective follow-up investigation." Frontiers in Medicine 9.

Ali, N. and K. Hossain (2020). "Liver injury in severe COVID-19 infection: current insights and challenges." Expert Review of Gastroenterology & Hepatology 14(10): 879–884.

Almulla, A. F., M. Maes, Z. Bo, K. A.-H. Hussein and V. Aristo (2023). "Brain-targeted autoimmunity is strongly associated with Long COVID and its chronic fatigue syndrome as well as its affective symptoms." medRxiv: 2023.2010.2004.23296554.

Almulla, A. F., Y. Thipakorn, A. A. A. Algon, C. Tunvirachaisakul, H. K. Al-Hakeim and M. Maes (2023). "Reverse cholesterol transport and lipid peroxidation biomarkers in major depression and bipolar disorder: A systematic review and meta-analysis." Brain, Behavior, and Immunity 113: 374–388.

Almulla, A. F., Y. Thipakorn, A. Vasupanrajit, A. A. Abo Algon, C. Tunvirachaisakul, A. A. Hashim Aljanabi, G. Oxenkrug, H. K. Al-Hakeim and M. Maes (2022a). "The tryptophan catabolite or kynurenine pathway in major depressive and bipolar disorder: A systematic review and meta-analysis." Brain, Behavior, & Immunity - Health 26: 100537.

Almulla, A. F., Y. Thipakorn, B. Zhou, A. Vojdani and M. Maes (2024). "Immune activation and immune-associated neurotoxicity in Long-COVID: A systematic review and meta-analysis of 103 studies comprising 58 cytokines/chemokines/growth factors." Brain, Behavior, and Immunity 122: 75–94.

Almulla, A. F., Y. Thipakorn, B. Zhou, A. Vojdani, R. Paunova and M. Maes (2024). "The tryptophan catabolite or kynurenine pathway in Long COVID disease: A systematic review and meta-analysis." medRxiv: 2024.2004.2030.24306635.

Andrés-Rodríguez, L., X. Borràs, A. Feliu-Soler, A. Pérez-Aranda, N. Angarita-Osorio, P. Moreno-Peral, J. Montero-Marin, J. García-Campayo, A. F. Carvalho, M. Maes and J. V. Luciano (2020). "Peripheral immune aberrations in fibromyalgia: A systematic review, meta-analysis and meta-regression." Brain, Behavior, and Immunity 87: 881–889.

Aparisi, Á., C. Ybarra-Falcón, M. García-Gómez, J. Tobar, C. Iglesias-Echeverría, S. Jaurrieta-Largo, R. Ladrón, A. Uribarri, P. Catalá, W. Hinojosa, M. Marcos-Mangas, L. Fernández-Prieto, R. Sedano-Gutiérrez, I. Cusacovich, D. Andaluz-Ojeda, B. de Vega-Sánchez, A. Recio-Platero, E. Sanz-Patiño, D. Calvo, C. Baladrón, M. Carrasco-Moraleja, C. Disdier-Vicente, I. J. Amat-Santos and J. A. San Román (2021). "Exercise Ventilatory Inefficiency in Post-COVID-19 Syndrome: Insights from a Prospective Evaluation." J Clin Med 10(12).

Aparisi, Á., C. Ybarra-Falcón, C. Iglesias-Echeverría, M. García-Gómez, M. Marcos-Mangas, G. Valle-Peñacoba, M. Carrasco-Moraleja, C. Fernández-de-Las-Peñas, L. Guerrero Á and D. García-Azorín (2022). "Cardio-Pulmonary Dysfunction Evaluation in Patients with Persistent Post-COVID-19 Headache." Int J Environ Res Public Health 19(7).

Aziz, M., R. Fatima, W. Lee-Smith and R. Assaly (2020). "The association of low serum albumin level with severe COVID-19: a systematic review and meta-analysis." Critical Care 24(1): 255.

Belenichev, I., L. Kucherenko, S. Pavlov, N. Bukhtiyarova, O. Popazova, N. Derevianko and G. Nimenko (2022). Therapy of post-COVID-19 syndrome: Improving the efficiency and safety of basic metabolic drug treatment with tiazotic acid (thiotriazoline). Pharmacia. 2022; 69 (2): 509–16.

Boleslav, L., T. Serdyukova and V. Skvortsov (2024). "Liver pathology in COVID-19 after end of pandemic: Modern view of problem." Medical alphabet: 10–15.

Boregowda, U., M. M. Aloysius, A. Perisetti, M. Gajendran, P. Bansal and H. Goyal (2020). "Serum Activity of Liver Enzymes Is Associated With Higher Mortality in COVID-19: A Systematic Review and Meta-Analysis." Front Med (Lausanne) 7: 431.

Boruga, M., S. Septimiu-Radu, P. S. Nandarge, A. Elagez, G. Doros, V. E. Lazureanu, E. R. Stoicescu, E. Tanase, R. Iacob, A. Dumitrescu, A. V. Bota, C. Cotoraci and M. L. Bratu (2024). "Kidney Function Tests and Continuous eGFR Decrease at Six Months after SARS-CoV-2 Infection in Patients Clinically Diagnosed with Post-COVID Syndrome." Biomedicines 12(5).

Bota, A. V., F. Bratosin, S. S. S. Bandi, I. Bogdan, D. V. Razvan, A. O. Toma, M. F. Indries, A. N. Csep, C. Cotoraci, M. Prodan, F. Marc, F. Ignuta and I. Marincu (2024). "A Comparative Analysis of Liver Injury Markers in Post-COVID Syndrome among Elderly Patients: A Prospective Study." J Clin Med 13(4).

Caballero-Alvarado, J., C. Zavaleta Corvera, B. Merino Bacilio, C. Ruiz Caballero and K. Lozano-Peralta (2023). "Post-COVID cholangiopathy: A narrative review." Gastroenterologia y Hepatologia 46(6): 474–482.

Cervia-Hasler, C., S. C. Brüningk, T. Hoch, B. Fan, G. Muzio, R. C. Thompson, L. Ceglarek, R. Meledin, P. Westermann, M. Emmenegger, P. Taeschler, Y. Zurbuchen, M. Pons, D. Menges, T. Ballouz, S. Cervia-Hasler, S. Adamo, M. Merad, A. W. Charney, M. Puhan, P. Brodin, J. Nilsson, A. Aguzzi, M. E. Raeber, C. B. Messner, N. D. Beckmann, K. Borgwardt and O. Boyman (2024). "Persistent complement dysregulation with signs of thromboinflammation in active Long Covid." Science 383(6680): eadg7942.

Cezar, R., L. Kundura, S. André, C. Lozano, T. Vincent, L. Muller, J.-Y. Lefrant, C. Roger, P.-G. Claret, S. Duvnjak, P. Loubet, A. Sotto, T.-A. Tran, J. Estaquier and P. Corbeau (2024). "T4 apoptosis in the acute phase of SARS-CoV-2 infection predicts long COVID." Frontiers in Immunology 14.

Chandler, J., M. Cumpston, T. Li, M. J. Page and V. Welch (2019). "Cochrane handbook for systematic reviews of interventions." Hoboken: Wiley.

Clemente, I., G. Sinatti, A. Cirella, S. J. Santini and C. Balsano (2022) "Alteration of Inflammatory Parameters and Psychological Post-Traumatic Syndrome in Long-COVID Patients." International Journal of Environmental Research and Public Health 19 DOI: 10.3390/ijerph19127103.

Colarusso, C., A. Maglio, M. Terlizzi, C. Vitale, A. Molino, A. Pinto, A. Vatrella and R. Sorrentino (2021). "Post-COVID-19 Patients Who Develop Lung Fibrotic-like Changes Have Lower Circulating Levels of IFN-β but Higher Levels of IL-1α and TGF-β." Biomedicines 9(12).

Corrêa, H. L., L. A. Deus, T. B. Araújo, A. L. Reis, C. E. N. Amorim, A. B. Gadelha, R. L. Santos, F. S. Honorato, D. Motta-Santos, C. Tzanno-Martins, R. V. P. Neves and T. S. Rosa (2022). "Phosphate and IL-10 concentration as predictors of long-covid in hemodialysis patients: A Brazilian study." Frontiers in Immunology 13.

D’Mello, C. and M. G. Swain (2014). "Liver–brain interactions in inflammatory liver diseases: Implications for fatigue and mood disorders." Brain, Behavior, and Immunity 35: 9–20.

Dennis, A., D. J. Cuthbertson, D. Wootton, M. Crooks, M. Gabbay, N. Eichert, S. Mouchti, M. Pansini, A. Roca-Fernandez, H. Thomaides-Brears, M. Kelly, M. Robson, L. Hishmeh, E. Attree, M. Heightman, R. Banerjee and A. Banerjee (2023). "Multi-organ impairment and long COVID: a 1-year prospective, longitudinal cohort study." J R Soc Med 116(3): 97–112.

Dennis, A., M. Wamil, J. Alberts, J. Oben, D. J. Cuthbertson, D. Wootton, M. Crooks, M. Gabbay, M. Brady, L. Hishmeh, E. Attree, M. Heightman, R. Banerjee and A. Banerjee (2021). "Multiorgan impairment in low-risk individuals with post-COVID-19 syndrome: a prospective, community-based study." BMJ Open 11(3): e048391.

Di Gennaro, L., P. Valentini, S. Sorrentino, M. A. Ferretti, E. De Candia, M. Basso, S. Lancellotti, R. De Cristofaro, C. De Rose, F. Mariani, R. Morello, I. Lazzareschi, L. Sigfrid, D. Munblit and D. Buonsenso (2022). "Extended coagulation profile of children with Long Covid: a prospective study." Scientific Reports 12(1): 18392.

Díaz-Salazar, S., R. Navas, L. Sainz-Maza, P. Fierro, M. Maamar, A. Artime, H. Basterrechea, B. Petitta, S. Pini, J. M. Olmos, C. Ramos, E. Pariente and J. L. Hernández (2022). "Blood group O is associated with post-COVID-19 syndrome in outpatients with a low comorbidity index." Infect Dis (Lond) 54(12): 897–908.

Dudar, I., O. Loboda, І. Shifris and Y. Honchar (2023). "Post-COVID syndrome and cognitive dysfunction in patients treated with hemodialysis." Ukrainian Journal of Nephrology and Dialysis(4 (80)): 66–77.

Dugani, P., A. Mehta, S. Furtado, R. Pradeep, M. Javali, P. Acharya, V. Thyagaraj and R. Srinivasa (2022). "Spectrum of neurological manifestations among acute COVID-19 and long COVID-19 – A retrospective observational study." Romanian Journal of Neurology 21: 176–182.

Effenberger, M., C. Grander, F. Grabherr, A. Griesmacher, T. Ploner, F. Hartig, R. Bellmann-Weiler, M. Joannidis, H. Zoller, G. Weiss, T. E. Adolph and H. Tilg (2021). "Systemic inflammation as fuel for acute liver injury in COVID-19." Dig Liver Dis 53(2): 158–165.

Ewing, A. G., S. Salamon, E. Pretorius, D. Joffe, G. Fox, S. Bilodeau and Y. Bar-Yam (2024). "Review of organ damage from COVID and Long COVID: a disease with a spectrum of pathology."

Fan, B. E., S. W. Wong, C. L. L. Sum, G. H. Lim, B. P. Leung, C. W. Tan, K. Ramanathan, R. Dalan, C. Cheung, X. R. Lim, M. S. Sadasiv, D. C. Lye, B. E. Young, E. S. Yap and Y. W. Chia (2022). "Hypercoagulability, endotheliopathy, and inflammation approximating 1 year after recovery: Assessing the long-term outcomes in COVID-19 patients." Am J Hematol 97(7): 915–923.

Fernandez-de-las-Peñas, C., K. I. Notarte, R. Macasaet, J. V. Velasco, J. A. Catahay, A. T. Ver, W. Chung, J. A. Valera-Calero and M. Navarro-Santana (2024). "Persistence of post-COVID symptoms in the general population two years after SARS-CoV-2 infection: A systematic review and meta-analysis." Journal of Infection 88(2): 77–88.

Fernández-de-Las-Peñas, C., P. Ryan-Murua, J. Rodríguez-Jiménez, M. Palacios-Ceña, L. Arendt-Nielsen and J. Torres-Macho (2022). "Serological Biomarkers at Hospital Admission Are Not Related to Long-Term Post-COVID Fatigue and Dyspnea in COVID-19 Survivors." Respiration 101(7): 658–665.

Frontera, J. A., R. A. Betensky, L. A. Pirofski, T. Wisniewski, H. Yoon and M. B. Ortigoza (2024). "Trajectories of Inflammatory Markers and Post-COVID-19 Cognitive Symptoms: A Secondary Analysis of the CONTAIN COVID-19 Randomized Trial." Neurol Neuroimmunol Neuroinflamm 11(3): e200227.

Gameil, M. A., R. E. Marzouk, A. H. Elsebaie and S. E. Rozaik (2021). "Long-term clinical and biochemical residue after COVID-19 recovery." Egyptian Liver Journal 11(1): 74.

García-Abellán, J., M. Fernández, S. Padilla, J. A. García, V. Agulló, V. Lozano, N. Ena, L. García-Sánchez, F. Gutiérrez and M. Masiá (2022). "Immunologic phenotype of patients with long-COVID syndrome of 1-year duration." Front Immunol 13: 920627.

Garcia-Gasalla, M., M. Berman-Riu, A. Rodriguez, A. Iglesias, P. A. Fraile-Ribot, N. Toledo-Pons, E. Pol-Pol, A. Ferré-Beltrán, F. Artigues-Serra, M. L. Martin-Pena, J. Pons, J. Murillas, A. Oliver, M. Riera and J. M. Ferrer (2023). "Elevated complement C3 and increased CD8 and type 1 helper lymphocyte T populations in patients with post-COVID-19 condition." Cytokine 169: 156295.

Goeijenbier, M., M. van Wissen, C. van de Weg, E. Jong, V. E. Gerdes, J. C. Meijers, D. P. Brandjes and E. C. van Gorp (2012). "Review: Viral infections and mechanisms of thrombosis and bleeding." J Med Virol 84(10): 1680–1696.

Gu, X., X. Li, X. An, S. Yang, S. Wu, X. Yang and H. Wang (2020). "Elevated serum aspartate aminotransferase level identifies patients with coronavirus disease 2019 and predicts the length of hospital stay." J Clin Lab Anal 34(7): e23391.

Guntur, V. P., T. Nemkov, E. de Boer, M. P. Mohning, D. Baraghoshi, F. I. Cendali, I. San-Millán, I. Petrache and A. D’Alessandro (2022). "Signatures of Mitochondrial Dysfunction and Impaired Fatty Acid Metabolism in Plasma of Patients with Post-Acute Sequelae of COVID-19 (PASC)." Metabolites 12(11).

Gupta, A., R. Nicholas, J. J. McGing, A. V. Nixon, J. E. Mallinson, T. M. McKeever, C. R. Bradley, M. Piasecki, E. F. Cox, J. Bonnington, J. M. Lord, C. E. Brightling, R. A. Evans, I. P. Hall, S. T. Francis, P. L. Greenhaff and C. E. Bolton (2024). "DYNamic Assessment of Multi-Organ level dysfunction in patients recovering from COVID-19: DYNAMO COVID-19." Exp Physiol 109(8): 1274–1291.

Hossain, A. A., I. U. Ahamed, U. D. Gupta, A. N. Anika and I. U. Ahamed (2024). Stratified Prognostication and Interventional Strategies in Chronic Hepatic Diseases: An Ensemble Machine Learning Approach. Proceedings - IEEE International Conference on Advanced Systems and Emergent Technologies, IC_ASET 2024.

Hu, X., L. Sun, Z. Guo, C. Wu, X. Yu and J. Li (2022). "Management of COVID-19 patients with chronic liver diseases and liver transplants: COVID-19 and liver diseases." Annals of Hepatology 27(1).

Huang, T.-L. and C.-T. Lee (2007). "Low serum albumin and high ferritin levels in chronichemodialysis patients with major depression." Psychiatry Research 152(2): 277–280.

Ishikura, T., T. Nakano, T. Kitano, T. Tokuda, H. Sumi-Akamaru and T. Naka (2023). "Serum ferritin level during hospitalization is associated with Brain Fog after COVID-19." Scientific Reports 13(1): 13095.

Ivchenko, G. S., N. N. Lobzhanidze, D. S. Rusina, E. V. Denisova and A. A. Ivchenko (2023). "[Mild post-COVID syndrome in young patients]." Ter Arkh 95(8): 674–678.

Kalinskaya, A., D. Vorobyeva, G. Rusakovich, E. Maryukhnich, A. Anisimova, O. Dukhin, A. Elizarova, O. Ivanova, A. Bugrova, A. Brzhozovskiy, A. Kononikhin, E. Nikolaev and E. Vasilieva (2023). "Targeted Blood Plasma Proteomics and Hemostasis Assessment of Post COVID-19 Patients with Acute Myocardial Infarction." Int J Mol Sci 24(7).

Kankaya, S., F. Yavuz, A. Tari, A. B. Aygun, E. G. Gunes, B. Bektan Kanat, G. Ulugerger Avci, H. Yavuzer and Y. Dincer (2023). "Glutathione-related antioxidant defence, DNA damage, and DNA repair in patients suffering from post-COVID conditions." Mutagenesis 38(4): 216–226.

Kerget, B., E. Çelik, F. Kerget, A. Aksakal, E. Y. Uçar, Ö. Araz and M. Akgün (2022). "Evaluation of 3-month follow-up of patients with postacute COVID-19 syndrome." J Med Virol 94(5): 2026–2034.

Kolesova, O., I. Vanaga, S. Laivacuma, A. Derovs, A. Kolesovs, M. Radzina, A. Platkajis, J. Eglite, E. Hagina, S. Arutjunana, D. S. Putrins, J. Storozenko, B. Rozentale and L. Viksna (2021). "Intriguing findings of liver fibrosis following COVID-19." BMC Gastroenterol 21(1): 370.

Korsunskiy, E. S., E. A. Belousova and A. A. Budzinskaya (2023). "The post-COVID-19 intestinal damages: clinical, endoscopic and morphological features. The results of a single-center prospective observational cohort study." Almanac of Clinical Medicine 51(8): 427–440.

Kovarik, J. J., A. Bileck, G. Hagn, S. M. Meier-Menches, T. Frey, A. Kaempf, M. Hollenstein, T. Shoumariyeh, L. Skos, B. Reiter, M. C. Gerner, A. Spannbauer, E. Hasimbegovic, D. Schmidl, G. Garhöfer, M. Gyöngyösi, K. G. Schmetterer and C. Gerner (2023). "A multi-omics based anti-inflammatory immune signature characterizes long COVID-19 syndrome." iScience 26(1): 105717.

Kruger, A., M. Vlok, S. Turner, C. Venter, G. J. Laubscher, D. B. Kell and E. Pretorius (2022). "Proteomics of fibrin amyloid microclots in long COVID/post-acute sequelae of COVID-19 (PASC) shows many entrapped pro-inflammatory molecules that may also contribute to a failed fibrinolytic system." Cardiovasc Diabetol 21(1): 190.

Kuchler, T., R. Günthner, A. Ribeiro, R. Hausinger, L. Streese, A. Wöhnl, V. Kesseler, J. Negele, T. Assali, J. Carbajo-Lozoya, M. Lech, H. Schneider, K. Adorjan, H. C. Stubbe, H. Hanssen, K. Kotilar, B. Haller, U. Heemann and C. Schmaderer (2023). "Persistent endothelial dysfunction in post-COVID-19 syndrome and its associations with symptom severity and chronic inflammation." Angiogenesis 26(4): 547–563.

Lazebnik, L., S. Turkina, R. Myazin, L. Tarasova, T. Ermolova, S. Kozhevnikova and D. Abdulganieva (2023). "Hyperammonemia as a manifestation of post-covid syndrome in patients with nonalcoholic fatty liver disease: post-hoc analysis of the LIRA - COVID observational clinical program." Experimental and Clinical Gastroenterology: 140–147.

Lebbe, A., A. Aboulwafa, N. Bayraktar, B. Mushannen, S. Ayoub, S. Sarker, M. N. Abdalla, I. Mohammed, M. Mushannen, L. Yagan and D. Zakaria (2024) "New Onset of Acute and Chronic Hepatic Diseases Post-COVID-19 Infection: A Systematic Review." Biomedicines 12 DOI: 10.3390/biomedicines12092065.

Lechuga, G. C., C. M. Morel and S. G. De-Simone (2023). "Hematological alterations associated with long COVID-19." Frontiers in Physiology 14.

Lee, Y. R., M. K. Kang, J. E. Song, H. J. Kim, Y. O. Kweon, W. Y. Tak, S. Y. Jang, J. G. Park, C. Lee, J. S. Hwang, B. K. Jang, J. I. Suh, W. J. Chung, B. S. Kim and S. Y. Park (2020). "Clinical outcomes of coronavirus disease 2019 in patients with pre-existing liver diseases: A multicenter study in South Korea." Clin Mol Hepatol 26(4): 562–576.

Levi, M., J. Thachil, T. Iba and J. H. Levy (2020). "Coagulation abnormalities and thrombosis in patients with COVID-19." The Lancet Haematology 7(6): e438–e440.

Li, X., C. Fan, J. Tang and N. Zhang (2023). "Meta-analysis of liver injury in patients with COVID-19." Medicine (Baltimore) 102(29): e34320.

Lim, J. S., J. H. Yang, B. Y. Chun, S. Kam, D. R. Jacobs Jr and D. H. Lee (2004). "Is serum γ-glutamyltransferase inversely associated with serum antioxidants as a marker of oxidative stress?" Free Radical Biology and Medicine 37(7): 1018–1023.

Lopez-Leon, S., T. Wegman-Ostrosky, C. Perelman, R. Sepulveda, P. A. Rebolledo, A. Cuapio and S. Villapol (2021). "More than 50 long-term effects of COVID-19: a systematic review and meta-analysis." Scientific Reports 11(1): 16144.

Lyalyukova, E., I. Dolgalev, E. Chernysheva, I. Druk, G. Konovalova and A. Lyalyukov (2021). "Liver damage while Covid-19: problems of pathogenesis and treatment."

Maamar, M., A. Artime, E. Pariente, P. Fierro, Y. Ruiz, S. Gutiérrez, M. Tobalina, S. Díaz-Salazar, C. Ramos, J. M. Olmos and J. L. Hernández (2022). "Post-COVID-19 syndrome, low-grade inflammation and inflammatory markers: a cross-sectional study." Curr Med Res Opin 38(6): 901–909.

Maes, M., H. T. Al-Rubaye, A. F. Almulla, D. S. Al-Hadrawi, K. Stoyanova, M. Kubera and H. K. Al-Hakeim (2022) "Lowered Quality of Life in Long COVID Is Predicted by Affective Symptoms, Chronic Fatigue Syndrome, Inflammation and Neuroimmunotoxic Pathways." International Journal of Environmental Research and Public Health 19 DOI: 10.3390/ijerph191610362.

Maes, M., A. F. Almulla, X. Tang, K. Stoyanova and A. Vojdani (2024). "From human herpes virus-6 reactivation to autoimmune reactivity against tight junctions and neuronal antigens, to inflammation, depression, and chronic fatigue syndrome due to Long COVID." Journal of Medical Virology 96(8): e29864.

Maes, M., J. Van de Vyvere, E. Vandoolaeghe, T. Bril, P. Demedts, A. Wauters and H. Neels (1996). "Alterations in iron metabolism and the erythron in major depression: further evidence for a chronic inflammatory process." J Affect Disord 40(1-2): 23–33.

Maes, M., M. Vandewoude, S. Scharpé, L. De Clercq, W. Stevens, L. Lepoutre and C. Schotte (1991). "Anthropometric and biochemical assessment of the nutritional state in depression: evidence for lower visceral protein plasma levels in depression." J Affect Disord 23(1): 25–33.

Magdy, R., R. A. Eid, W. Fathy, M. M. Abdel-Aziz, R. E. Ibrahim, A. Yehia, M. S. Sheemy and M. Hussein (2022). "Characteristics and Risk Factors of Persistent Neuropathic Pain in Recovered COVID-19 Patients." Pain Med 23(4): 774–781.

Malakouti, M., A. Kataria, S. K. Ali and S. Schenker (2017). "Elevated Liver Enzymes in Asymptomatic Patients - What Should I Do?" J Clin Transl Hepatol 5(4): 394–403.

Marín-Gómez, L. M., C. Bernal-Bellido, J. M. Alamo-Martínez, F. M. Porras-López, G. Suárez-Artacho, J. Serrano-Diaz-Canedo, J. Padillo-Ruiz and M. A. Gómez-Bravo (2012). "Intraoperative hepatic artery blood flow predicts early hepatic artery thrombosis after liver transplantation." Transplant Proc 44(7): 2078–2081.

Martone, A. M., M. Tosato, F. Ciciarello, V. Galluzzo, M. B. Zazzara, C. Pais, G. Savera, R. Calvani, E. Marzetti, M. C. Robles, M. Ramirez and F. Landi (2022). "Sarcopenia as potential biological substrate of long COVID-19 syndrome: prevalence, clinical features, and risk factors." J Cachexia Sarcopenia Muscle 13(4): 1974–1982.

Meisinger, C., Y. Goßlau, T. D. Warm, V. Leone, A. Hyhlik-Dürr, J. Linseisen and I. Kirchberger (2022). "Post-COVID-19 Fatigue and SARS-CoV-2 Specific Humoral and T-Cell Responses in Male and Female Outpatients." Frontiers in Immunology 13.

Nádasdi, Á., G. Sinkovits, I. Bobek, B. Lakatos, Z. Förhécz, Z. Z. Prohászka, M. Réti, M. Arató, G. Cseh, T. Masszi, B. Merkely, P. Ferdinandy, I. Vályi-Nagy, Z. Prohászka and G. Firneisz (2022). "Decreased circulating dipeptidyl peptidase-4 enzyme activity is prognostic for severe outcomes in COVID-19 inpatients." Biomark Med 16(5): 317–330.

Nasir, N., I. Khanum, K. Habib, A. Wagley, A. Arshad and A. Majeed (2024). "Insight into COVID-19 associated liver injury: Mechanisms, evaluation, and clinical implications." Hepatol Forum 5(3): 139–149.

Negrut, N., G. Menegas, S. Kampioti, M. Bourelou, F. Kopanyi, F. D. Hassan, A. Asowed, F. Z. Taleouine, A. Ferician and P. Marian (2024). "The Multisystem Impact of Long COVID: A Comprehensive Review." Diagnostics (Basel) 14(3).

Ngiam, J. N., N. Chew, S. M. Tham, Z. Y. Lim, T. Y. Li, S. Cen, P. A. Tambyah, A. Santosa, M. Muthiah, C. H. Sia and G. B. Cross (2021). "Elevated liver enzymes in hospitalized patients with COVID-19 in Singapore." Medicine (Baltimore) 100(30): e26719.

Nicolai, L., R. Kaiser and K. Stark (2023). "Thromboinflammation in long COVID-the elusive key to postinfection sequelae?" J Thromb Haemost 21(8): 2020–2031.

Nivukoski, U., A. Bloigu, R. Bloigu, M. Aalto, T. Laatikainen and O. Niemelä (2019). "Liver enzymes in alcohol consumers with or without binge drinking." Alcohol 78: 13–19.

Page, M. J., J. E. McKenzie, P. M. Bossuyt, I. Boutron, T. C. Hoffmann, C. D. Mulrow, L. Shamseer, J. M. Tetzlaff, E. A. Akl and S. E. Brennan (2021). "The PRISMA 2020 statement: an updated guideline for reporting systematic reviews." bmj 372.

Pan, B., X. Wang, H. Lai, R. W. M. Vernooij, X. Deng, N. Ma, D. Li, J. Huang, W. Zhao, J. Ning, J. Liu, J. Tian, L. Ge and K. Yang (2024). "Risk of kidney and liver diseases after COVID-19 infection: A systematic review and meta-analysis." Rev Med Virol 34(2): e2523.

Paniskaki, K., S. Goretzki, M. Anft, M. J. Konik, T. L. Meister, S. Pfaender, K. Lechtenberg, M. Vogl, B. Dogan, S. Dolff, T. H. Westhoff, H. Rohn, U. Felderhoff-Mueser, U. Stervbo, O. Witzke, C. Dohna-Schwake and N. Babel (2023). "Increased SARS-CoV-2 reactive low avidity T cells producing inflammatory cytokines in pediatric post-acute COVID-19 sequelae (PASC)." Pediatr Allergy Immunol 34(12): e14060.

Papadopoulos, N., S. Vasileiadi and M. Deutsch (2020). "COVID-19 and liver injury: Where do we stand?" Annals of Gastroenterology 33(5): 459–464.

Parás-Bravo, P., C. Fernández-de-las-Peñas, D. Ferrer-Pargada, S. Izquierdo-Cuervo, L. M. Fernández-Cacho, J. M. Cifrián-Martínez, P. Druet-Toquero, O. Pellicer-Valero and M. Herrero-Montes (2024) "Serological Biomarkers in Individuals with Interstitial Lung Disease after SARS-CoV-2 Infection and Association with Post-COVID-19 Symptoms." Pathogens 13 DOI: 10.3390/pathogens13080641.

Paris, D., L. Palomba, M. C. Albertini, A. Tramice, L. Motta, E. Giammattei, P. Ambrosino, M. Maniscalco and A. Motta (2023). "The biomarkers’ landscape of post-COVID-19 patients can suggest selective clinical interventions." Sci Rep 13(1): 22496.

Pasini, E., G. Corsetti, C. Romano, T. M. Scarabelli, C. Chen-Scarabelli, L. Saravolatz and F. S. Dioguardi (2021). "Serum Metabolic Profile in Patients With Long-Covid (PASC) Syndrome: Clinical Implications." Front Med (Lausanne) 8: 714426.

Petramala, L., F. Sarlo, A. Servello, S. Baroni, M. Suppa, F. Circosta, G. Galardo, O. Gandini, L. Marino, G. Cavallaro, G. Iannucci, A. Concistrè and C. Letizia (2023). "Pulmonary embolism post-Covid-19 infection: physiopathological mechanisms and vascular damage biomarkers." Clin Exp Med 23(8): 4871–4880.

Portacci, A., M. Amendolara, V. N. Quaranta, I. Iorillo, E. Buonamico, F. Diaferia, S. Quaranta, C. Locorotondo, A. Schirinzi, E. Boniello, S. Dragonieri and G. E. Carpagnano (2024). "Can Galectin-3 be a reliable predictive biomarker for post-COVID syndrome development?" Respir Med 226: 107628.

Portugal-Nunes, C., T. C. Castanho, L. Amorim, P. S. Moreira, J. Mariz, F. Marques, N. Sousa, N. C. Santos and J. A. Palha (2020). "Iron Status is Associated with Mood, Cognition, and Functional Ability in Older Adults: A Cross-Sectional Study." Nutrients 12(11).

Poyatos, P., N. Luque, G. Sabater, S. Eizaguirre, M. Bonnin, R. Orriols and O. Tura-Ceide (2024). "Endothelial dysfunction and cardiovascular risk in post-COVID-19 patients after 6- and 12-months SARS-CoV-2 infection." Infection.

Pretorius, E., M. Vlok, C. Venter, J. A. Bezuidenhout, G. J. Laubscher, J. Steenkamp and D. B. Kell (2021). "Persistent clotting protein pathology in Long COVID/Post-Acute Sequelae of COVID-19 (PASC) is accompanied by increased levels of antiplasmin." Cardiovascular Diabetology 20(1): 172.

Radzina, M., D. S. Putrins, A. Micena, I. Vanaga, O. Kolesova, A. Platkajis and L. Viksna (2022). "Post-COVID-19 Liver Injury: Comprehensive Imaging With Multiparametric Ultrasound." J Ultrasound Med 41(4): 935–949.

Raman, B., M. P. Cassar, E. M. Tunnicliffe, N. Filippini, L. Griffanti, F. Alfaro-Almagro, T. Okell, F. Sheerin, C. Xie, M. Mahmod, F. E. Mózes, A. J. Lewandowski, E. O. Ohuma, D. Holdsworth, H. Lamlum, M. J. Woodman, C. Krasopoulos, R. Mills, F. A. K. McConnell, C. Wang, C. Arthofer, F. J. Lange, J. Andersson, M. Jenkinson, C. Antoniades, K. M. Channon, M. Shanmuganathan, V. M. Ferreira, S. K. Piechnik, P. Klenerman, C. Brightling, N. P. Talbot, N. Petousi, N. M. Rahman, L. P. Ho, K. Saunders, J. R. Geddes, P. J. Harrison, K. Pattinson, M. J. Rowland, B. J. Angus, F. Gleeson, M. Pavlides, I. Koychev, K. L. Miller, C. Mackay, P. Jezzard, S. M. Smith and S. Neubauer (2021). "Medium-term effects of SARS-CoV-2 infection on multiple vital organs, exercise capacity, cognition, quality of life and mental health, post-hospital discharge." EClinicalMedicine 31: 100683.

Ren, M., C. Lu, M. Zhou, X. Jiang, X. Li and N. Liu (2024). "The intersection of virus infection and liver disease: A comprehensive review of pathogenesis, diagnosis, and treatment." WIREs Mech Dis 16(3): e1640.

Rinaldi, L., S. Rigo, M. Pani, A. Bisoglio, K. Khalaf, M. Minonzio, D. Shiffer, M. A. Romeo, P. Verzeletti, M. Ciccarelli, M. G. Bordoni, S. Stranges, E. Riboli, R. Furlan and F. Barbic (2024). "Long-COVID autonomic syndrome in working age and work ability impairment." Scientific Reports 14(1): 11835.

Sasso, E. M., K. Muraki, N. Eaton-Fitch, P. Smith, O. L. Lesslar, G. Deed and S. Marshall-Gradisnik (2022). "Transient receptor potential melastatin 3 dysfunction in post COVID-19 condition and myalgic encephalomyelitis/chronic fatigue syndrome patients." Mol Med 28(1): 98.

Savage, K., C. Porter, E. Bunnett, M. Hana, A. Keegan, E. Ogden, C. Stough and A. Pipingas (2023). "Liver and inflammatory biomarker relationships to depression symptoms in healthy older adults." Exp Gerontol 177: 112186.

Shi, Y., M. Wang, L. Wu, X. Li and Z. Liao (2023). "COVID-19 associated liver injury: An updated review on the mechanisms and management of risk groups." Liver Research 7(3): 207–215.

Sibila, O., L. Perea, N. Albacar, J. Moisés, T. Cruz, N. Mendoza, B. Solarat, G. Lledó, G. Espinosa, J. A. Barberà, J. R. Badia, A. Agustí, J. Sellarés and R. Faner (2022). "Elevated plasma levels of epithelial and endothelial cell markers in COVID-19 survivors with reduced lung diffusing capacity six months after hospital discharge." Respiratory Research 23(1): 37.

Simental-Mendía, L. E., M. Rodríguez-Morán, R. Gómez-Díaz, N. H. Wacher, H. Rodríguez-Hernández and F. Guerrero-Romero (2017). "Insulin resistance is associated with elevated transaminases and low aspartate aminotransferase/alanine aminotransferase ratio in young adults with normal weight." European Journal of Gastroenterology & Hepatology 29(4).

Sommen, S. L., L. B. Havdal, J. Selvakumar, G. Einvik, T. M. Leegaard, F. Lund-Johansen, A. E. Michelsen, T. E. Mollnes, T. Stiansen-Sonerud, T. Tjade, V. B. B. Wyller and L. L. Berven (2023). "Inflammatory markers and pulmonary function in adolescents and young adults 6 months after mild COVID-19." Frontiers in Immunology 13.

Stavileci, B., E. Özdemir, B. Özdemir, E. Ereren and M. Cengiz (2022). "De-novo development of fragmented QRS during a six-month follow-up period in patients with COVID-19 disease and its cardiac effects." J Electrocardiol 72: 44–48.

Stufano, A., C. Isgrò, L. L. Palese, P. Caretta, L. De Maria, P. Lovreglio and A. M. Sardanelli (2023). "Oxidative Damage and Post-COVID Syndrome: A Cross-Sectional Study in a Cohort of Italian Workers." Int J Mol Sci 24(8).

Sumbalova, Z., J. Kucharska, P. Palacka, Z. Rausova, P. H. Langsjoen, A. M. Langsjoen and A. Gvozdjakova (2022). "Platelet mitochondrial function and endogenous coenzyme Q10 levels are reduced in patients after COVID-19." Bratisl Lek Listy 123(1): 9–15.

Sumbalová, Z., J. Kucharská, Z. Rausová, P. Palacka, E. Kovalčíková, T. Takácsová, V. Mojto, P. Navas, G. Lopéz-Lluch and A. Gvozdjáková (2022). "Reduced platelet mitochondrial respiration and oxidative phosphorylation in patients with post COVID-19 syndrome are regenerated after spa rehabilitation and targeted ubiquinol therapy." Frontiers in Molecular Biosciences 9.

Sunada, N., H. Honda, Y. Nakano, K. Yamamoto, K. Tokumasu, Y. Sakurada, Y. Matsuda, T. Hasegawa, Y. Otsuka, M. Obika, Y. Hanayama, H. Hagiya, K. Ueda, H. Kataoka and F. Otsuka (2022). "Hormonal trends in patients suffering from long COVID symptoms." Endocr J 69(10): 1173–1181.

Taha, S. I., S. F. Samaan, R. A. Ibrahim, E. M. El-Sehsah and M. K. Youssef (2021). "Post-COVID-19 arthritis: is it hyperinflammation or autoimmunity?" Eur Cytokine Netw 32(4): 83–88.

Talla, A., S. V. Vasaikar, G. L. Szeto, M. P. Lemos, J. L. Czartoski, H. MacMillan, Z. Moodie, K. W. Cohen, L. B. Fleming, Z. Thomson, L. Okada, L. A. Becker, E. M. Coffey, S. C. De Rosa, E. W. Newell, P. J. Skene, X. Li, T. F. Bumol, M. J. McElrath and T. R. Torgerson (2022). "Persistent serum protein signatures define an inflammatory subset of long COVID." bioRxiv: 2022.2005.2009.491196.

Teng, L., X. Song, M. Zhang, Y. Han, G. Chang, W. Chang and Z. Shen (2023). "The pattern of cytokines expression and dynamic changes of renal function at 6 months in patients with Omicron COVID-19." J Med Virol 95(2): e28477.

Torki, E., F. Hoseininasab, M. Moradi, R. Sami, M. J. M. Sullman and H. Fouladseresht (2024). "The demographic, laboratory and genetic factors associated with long Covid-19 syndrome: a case-control study." Clin Exp Med 24(1): 1.

Townsend, L., A. H. Dyer, K. Jones, J. Dunne, A. Mooney, F. Gaffney, L. O’Connor, D. Leavy, K. O’Brien, J. Dowds, J. A. Sugrue, D. Hopkins, I. Martin-Loeches, C. Ni Cheallaigh, P. Nadarajan, A. M. McLaughlin, N. M. Bourke, C. Bergin, C. O’Farrelly, C. Bannan and N. Conlon (2020). "Persistent fatigue following SARS-CoV-2 infection is common and independent of severity of initial infection." PLoS One 15(11): e0240784.

Turner, S., M. A. Khan, D. Putrino, A. Woodcock, D. B. Kell and E. Pretorius (2023). "Long COVID: pathophysiological factors and abnormalities of coagulation." Trends in Endocrinology & Metabolism 34(6): 321–344.

Vojdani, A., A. F. Almulla, B. Zhou, H. K. Al-Hakeim and M. Maes (2024). "Reactivation of herpesvirus type 6 and IgA/IgM-mediated responses to activin-A underpin long COVID, including affective symptoms and chronic fatigue syndrome." Acta Neuropsychiatr: 1–13.

Vollrath, S., L. Matits, A. Jerg, J. Zorn, L. John, J. M. Steinacker and D. A. Bizjak (2023). "Blood Profiling of Athletes after COVID-19: Differences in Blood Profiles of Post-COVID-19 Athletes Compared to Uninfected Athletic Individuals-An Exploratory Analysis." Biomedicines 11(7).

Walker, M., E. M. Federico, D. B. Kell, C. Dupont, A. Proal and E. Pretorius (2022). "Abstract 15637: Detection and Characterization of Fibrin/Amyloid Microclots in Patients With Post-Acute Sequelae of Covid-19." Circulation 146(Suppl_1): A15637–A15637.

Wallis, T. J. M., E. Heiden, J. Horno, B. Welham, H. Burke, A. Freeman, L. Dexter, A. Fazleen, A. Kong, C. McQuitty, M. Watson, S. Poole, N. J. Brendish, T. W. Clark, T. M. A. Wilkinson, M. G. Jones and B. G. Marshall (2021). "Risk factors for persistent abnormality on chest radiographs at 12-weeks post hospitalisation with PCR confirmed COVID-19." Respir Res 22(1): 157.

Wannamethee, S. G. and A. G. Shaper (2010). "Cigarette smoking and serum liver enzymes: the role of alcohol and inflammation." Ann Clin Biochem 47(Pt 4): 321–326.

Wijarnpreecha, K., P. Ungprasert, P. Panjawatanan, D. M. Harnois, H. B. Zaver, A. Ahmed and D. Kim (2021). "COVID-19 and liver injury: a meta-analysis." Eur J Gastroenterol Hepatol 33(7): 990–995.

World Health Organization, W. (2021). "A clinical case definition of post COVID-19 condition by a Delphi consensus 6 October." WHO.

World Health Organization, W. (2022). "A healthy return: Investment case for a sustainably financed WHO."

Wu, X., K. Q. Deng, C. Li, Z. Yang, H. Hu, H. Cai, C. Zhang, T. He, F. Zheng, H. Wang, X. A. Zhang, A. Caillon, Y. Yuan, X. Wang, H. Xu and Z. Lu (2021). "Cardiac Involvement in Recovered Patients From COVID-19: A Preliminary 6-Month Follow-Up Study." Front Cardiovasc Med 8: 654405.

Yamamoto, J. M., S. Prado-Núñez, M. Guarnizo-Poma, H. Lazaro-Alcantara, S. Paico-Palacios, B. Pantoja-Torres, V. del Carmen Ranilla-Seguin and V. A. Benites-Zapata (2020). "Association between serum transaminase levels and insulin resistance in euthyroid and non-diabetic adults." Diabetes & Metabolic Syndrome: Clinical Research & Reviews 14(1): 17–21.

Yamamoto, Y., Y. Otsuka, K. Tokumasu, N. Sunada, Y. Nakano, H. Honda, Y. Sakurada, T. Hasegawa, H. Hagiya and F. Otsuka (2023) "Utility of Serum Ferritin for Predicting Myalgic Encephalomyelitis/Chronic Fatigue Syndrome in Patients with Long COVID." Journal of Clinical Medicine 12 DOI: 10.3390/jcm12144737.

Yamamoto, Y., Y. Otsuka, K. Tokumasu, N. Sunada, Y. Nakano, H. Honda, Y. Sakurada, T. Hasegawa, H. Hagiya and F. Otsuka (2023). "Utility of Serum Ferritin for Predicting Myalgic Encephalomyelitis/Chronic Fatigue Syndrome in Patients with Long COVID." J Clin Med 12(14).

Yilmaz, N., E. Eren, C. Öz, Z. Kalayci and F. Saribek (2021). "Covid-19 and iron metabolism: Traditional review." Turkiye Klinikleri Journal of Medical Sciences 41(2): 176–188.

Yong, S. J., A. Halim, M. Halim, S. Liu, M. Aljeldah, B. R. Al Shammari, S. Alwarthan, M. Alhajri, A. Alawfi, A. Alshengeti, F. Khamis, J. Alsalman, A. N. Alshukairi, N. A. Abukhamis, F. S. Almaghrabi, S. A. Almuthree, A. M. Alsulaiman, B. M. Alshehail, A. H. Alfaraj, S. A. Alhawaj, R. K. Mohapatra and A. A. Rabaan (2023). "Inflammatory and vascular biomarkers in post-COVID-19 syndrome: A systematic review and meta-analysis of over 20 biomarkers." Reviews in Medical Virology 33(2): e2424.

Zelber-Sagi, S., S. Toker, G. Armon, S. Melamed, S. Berliner, I. Shapira, Z. Halpern, E. Santo and O. Shibolet (2013). "Elevated alanine aminotransferase independently predicts new onset of depression in employees undergoing health screening examinations." Psychological Medicine 43(12): 2603–2613.

