## supplementary file for "Elevated Liver Damage Biomarkers in Long COVID: A Systematic Review and Meta-Analysis"

**SHORT TITLE:** Liver Damage in Long COVID

(1-3) Abbas F. Almulla, (4) Yanin Thipakorn, (1,2,4,5-8) Michael Maes

1. Sichuan Provincial Center for Mental Health, Sichuan Provincial People's Hospital, School of Medicine, University of Electronic Science and Technology of China, Chengdu 610072, China

2. Key Laboratory of Psychosomatic Medicine, Chinese Academy of Medical Sciences, Chengdu 610072, China

3. Medical Laboratory Technology Department, College of Medical Technology, The Islamic University, Najaf, Iraq

4. Department of Psychiatry, Faculty of Medicine, Chulalongkorn University and King Chulalongkorn Memorial Hospital, the Thai Red Cross Society, Bangkok, Thailand.

5. Cognitive Fitness and Technology Research Unit, Faculty of Medicine, Chulalongkorn University, Bangkok, Thailand.

6. Department of Psychiatry, Medical University of Plovdiv, Plovdiv, Bulgaria.

7. Research Institute, Medical University Plovdiv, Plovdiv, Bulgaria.

8. Kyung Hee University, 26 Kyungheedae-ro, Dongdaemun-gu, Seoul 02447, Republic of Korea.

**Corresponding author:**

Prof. Dr. Michael Maes, M.D., Ph.D.

Sichuan Provincial Center for Mental Health

Sichuan Provincial People’s Hospital,

School of Medicine,

University of Electronic Science and Technology of China

Chengdu 610072

China

[Michael Maes Google Scholar profile](https://scholar.google.com/citations?user=1wzMZ7UAAAAJ&hl=en)

<https://scholar.google.co.th/citations?user=1wzMZ7UAAAAJ&hl=th&oi=ao>

Highly cited author: 2003-2023 (ISI, Clarivate)

ScholarGPS: Worldwide #1 in molecular neuroscience; #1/4 in pathophysiology

Expert worldwide medical expertise ranking, Expertscape (December 2022), worldwide:

#1 in CFS, #1 in oxidative stress, #1 in encephalomyelitis, #1 in nitrosative stress, #1 in nitrosation, #1 in tryptophan, #1 in aromatic amino acids, #1 in stress (physiological), #1 in neuroimmune; #2 in bacterial translocation; #3 in inflammation, #4-5: in depression, fatigue and psychiatry.

**ESF. Table 1.** Search sentences and terms were used in each database.

| **Database Name** | **Search Sentence** | **No. of Articles** |
| --- | --- | --- |
| **PubMed/Medline** | ("Long COVID" OR "Post-Acute COVID-19 Syndrome" OR "Post-COVID Syndrome") AND ("Liver Biomarkers" OR "Liver Enzymes" OR "Alanine Transaminase" OR "Aspartate Aminotransferase" OR "Alkaline Phosphatase" OR "Gamma-Glutamyltransferase" OR "Bilirubin" OR "Albumin") | **29** |
|  | ("Long COVID" OR "Post-Acute COVID-19 Syndrome" OR "Post-COVID Syndrome") AND ("Liver Dysfunction" OR "Liver Disease" OR "Liver Injury" OR "Hepatopathy") AND ("Biomarkers" OR "Liver Enzymes" OR "ALT" OR "AST" OR "Bilirubin" OR "Albumin") | **7** |
|  | ("Long COVID" OR "Post-Acute COVID-19 Syndrome" OR "Post-COVID Syndrome") AND ("Alanine Transaminase" OR "Aspartate Aminotransferase" OR "Alkaline Phosphatase" OR "Gamma-Glutamyltransferase" OR "Bilirubin" OR "Albumin" OR "Prothrombin Time" OR "Liver Enzymes") | **35** |
|  | ("Long COVID" OR "Post-Acute COVID-19 Syndrome" OR "Post-COVID Syndrome") AND ("Liver Dysfunction" OR "Liver Disease" OR "Liver Injury") AND ("Inflammation" OR "Cytokines" OR "C-Reactive Protein") | **6** |
| **Google Scholar** | "Long COVID" OR "Post-Acute COVID-19 Syndrome" OR "Post-COVID Syndrome" AND "Liver Dysfunction" OR "Liver Disease" OR "Liver Injury" OR "Hepatopathy" AND "Biomarkers" OR "Liver Enzymes" OR "ALT" OR "AST" OR "Bilirubin" OR "Albumin" | **3,970** |
|  | "Post-Acute COVID-19 Syndrome" OR "Long COVID" OR "Post-COVID Syndrome" AND "Alanine Transaminase" OR "Aspartate Aminotransferase" OR "Alkaline Phosphatase" OR "Gamma-Glutamyltransferase" OR "Bilirubin" OR "Albumin" OR "Prothrombin Time" OR "Liver Enzymes" | **14,800** |
|  | "Long COVID" OR "Post-Acute COVID-19 Syndrome" OR "Post-COVID Syndrome" AND "Liver Dysfunction" OR "Liver Disease" OR "Liver Injury" AND "Inflammation" OR "Cytokines" OR "C-Reactive Protein" | **4,630** |
| **SciFinder** | "Long COVID" OR "Post-Acute COVID-19 Syndrome" OR "Post-COVID Syndrome" AND "Liver Dysfunction" OR "Liver Disease" OR "Liver Injury" OR "Hepatopathy" AND "Biomarkers" OR "Liver Enzymes" OR "ALT" OR "AST" OR "Bilirubin" OR "Albumin" | **12** |
| **SCOPUS** | Long COVID" AND "Liver Function" OR "Liver Injury | **38** |

**ESF. Table 2.** Immune cofounder’s scale (ICS) applied from Andrés-Rodríguez. et al.. 2019

| **Methodological quality of the study** | |
| --- | --- |
| **1** | Study sample ≥ 128 participants including patients and controls (1= Yes. 0 = No) |
| **2** | Did the study control the results for potential confounders (e.g.. age. BMI. gender. race)? (1= Yes. 0 = No) |
| **3** | Were participants with Long COVID and controls age- and-gender-matched or was there a statistical control? (1= Yes. 0 = No) |
| **4** | Was the time of sample collection specified (e.g.. morning vs. evening)? (1= Yes. 0 = No) |
| **5** | Were participants with Long COVID free of immunomodulatory drugs including anti-cytokines. glucocorticoids. immunoglobulins. and immunosuppressants. or was there a medication washout period. or was drug intake statistically controlled for? (1= Yes. 0 = No) |
| **6** | Were participants with Long COVID free of antidepressants and mood stabilizers or were the data statistically controlled for? (1= Yes. 0 = No) |
| **7** | Reporting either the manufacturer of the test or detection limit and coefficients of variation (1= Yes. 0 = No) |
| **8** | Reporting how data under detection limit were handled (1 = Yes. 0 = No) |
| **9** | Reporting % of the sample under detection limit (1=Yes. 0= No) |
| **10** | Reporting blood fraction (serum. plasma. culture supernatant or whole blood) (1= Yes. 0 = No) |
| **Total quality score (10 points)** | |
| **Biomarker confounders red points**  *The red points should not be given if the item is statistically controlled for* | |
| **1** | 3 red points for comorbid illnesses such as autoimmune disorders & other immune disorders including rheumatoid arthritis. psoriasis. inflammatory bowel disease. chronic obstructive pulmonary disease. multiple sclerosis |
| **2** | 3 red points for use of recreational drugs such as methamphetamine or opioids |
| **3** | 2 red points when groups were not matched for age |
| **4** | 2 red points when groups were not matched for sex |
| **5** | 2 red points for medication use as for example immunomodulators |
| **6** | 2 red points for early traumatic life events |
| **7** | 2 red points for shift work and primary sleep disorders |
| **8** | 1.5 red points for use of antipsychotics |
| **9** | 1 red point for more common systemic immune disorders including diabetes type 1/2. essential hypertension. metabolic syndrome |
| **10** | 1 red point for not fasting (8 hours before blood extraction) |
| **11** | 1 red point for use of omega-3 and antioxidant supplements |
| **12** | 1 red point when data were not controlled for body mass index |
| **13** | 1 red point when data were not controlled for physical activity or sedentary life |
| **14** | 1 red point when data were not controlled for smoking |
| **15** | 1 red point for use of oral contraceptives or NSAIDs |
| **16** | 0.5 red points when data were not controlled for ethnicity in countries such as US. Brazil |
| **17** | 0.5 red points when data were not controlled for seasonality |
| **18** | 0.5 red points when data were not controlled for diurnal variation (8-10 a.m. versus all other time points) |
|  | **Total red point score (26 points)** |

**ESF. Table 3.** PRISMA checklist

| **Section/topic** | **#** | **Checklist item** | **Reported on page #** |
| --- | --- | --- | --- |
| **TITLE** | | | |
| Title | 1 | Identify the report as a systematic review. meta-analysis. or both. | 1 |
| **ABSTRACT** | | | |
| Structured summary | 2 | Provide a structured summary including. as applicable: background; objectives; data sources; study eligibility criteria. participants. and interventions; study appraisal and synthesis methods; results; limitations; conclusions and implications of key findings; systematic review registration number. | 3 |
| **INTRODUCTION** | | | |
| Rationale | 3 | Describe the rationale for the review in the context of what is already known. | 5 |
| Objectives | 4 | Provide an explicit statement of questions being addressed with reference to participants. interventions. comparisons. outcomes. and study design (PICOS). | 8 |
| **METHODS** | | | |
| Protocol and registration | 5 | Indicate if a review protocol exists. if and where it can be accessed (e.g.. Web address). and. if available. provide registration information including registration number. | 9 |
| Eligibility criteria | 6 | Specify study characteristics (e.g.. PICOS. length of follow-up) and report characteristics (e.g.. years considered. language. publication status) used as criteria for eligibility. giving rationale. | 10 |
| Information sources | 7 | Describe all information sources (e.g.. databases with dates of coverage. contact with study authors to identify additional studies) in the search and date last searched. | 9 |
| Search | 8 | Present full electronic search strategy for at least one database. including any limits used. such that it could be repeated. | ESF. Table 1. page 3 |
| Study selection | 9 | State the process for selecting studies (i.e.. screening. eligibility. included in systematic review. and. if applicable. included in the meta-analysis). | 10 |
| Data collection process | 10 | Describe method of data extraction from reports (e.g.. piloted forms. independently. in duplicate) and any processes for obtaining and confirming data from investigators. | 11 |
| Data items | 11 | List and define all variables for which data were sought (e.g.. PICOS. funding sources) and any assumptions and simplifications made. | 11 |
| Risk of bias in individual studies | 12 | Describe methods used for assessing risk of bias of individual studies (including specification of whether this was done at the study or outcome level). and how this information is to be used in any data synthesis. | 12 |
| Summary measures | 13 | State the principal summary measures (e.g.. risk ratio. difference in means). | 12 |
| Synthesis of results | 14 | Describe the methods of handling data and combining results of studies. if done. including measures of consistency (e.g.. I^2^) for each meta-analysis. | 12 |
| Risk of bias across studies | 15 | Specify any assessment of risk of bias that may affect the cumulative evidence (e.g.. publication bias. selective reporting within studies). | Table 3. page 71 |
| Additional analyses | 16 | Describe methods of additional analyses (e.g.. sensitivity or subgroup analyses. meta-regression). if done. indicating which were pre-specified. | 12 |
| **RESULTS** | | |  |
| Study selection | 17 | Give numbers of studies screened. assessed for eligibility. and included in the review. with reasons for exclusions at each stage. ideally with a flow diagram. | 14 |
| Study characteristics | 18 | For each study. present characteristics for which data were extracted (e.g.. study size. PICOS. follow-up period) and provide the citations. | ESF. Table 2 page |
| Risk of bias within studies | 19 | Present data on risk of bias of each study and. if available. any outcome level assessment (see item 12). | Table 3. page 71 |
| Results of individual studies | 20 | For all outcomes considered (benefits or harms). present. for each study: (a) simple summary data for each intervention group (b) effect estimates and confidence intervals. ideally with a forest plot. | Table 1. page 66 |
| Synthesis of results | 21 | Present results of each meta-analysis done. including confidence intervals and measures of consistency. | Table 2. page 68 |
| Risk of bias across studies | 22 | Present results of any assessment of risk of bias across studies (see Item 15). | Table 3 page 71 |
| Additional analysis | 23 | Give results of additional analyses. if done (e.g.. sensitivity or subgroup analyses. meta-regression [see Item 16]). | Table 2. page 68 |
| **DISCUSSION** | | |  |
| Summary of evidence | 24 | Summarize the main findings including the strength of evidence for each main outcome; consider their relevance to key groups (e.g.. healthcare providers. users. and policy makers). | Page 22-31 |
| Limitations | 25 | Discuss limitations at study and outcome level (e.g.. risk of bias). and at review-level (e.g.. incomplete retrieval of identified research. reporting bias). | Page 31-32 |
| Conclusions | 26 | Provide a general interpretation of the results in the context of other evidence. and implications for future research. | Page 32 |
| **FUNDING** | | |  |
| Funding | 27 | Describe sources of funding for the systematic review and other support (e.g.. supply of data); role of funders for the systematic review. | Page 33 |

**ESF. table 4.** Characteristics of the studies included in the systematic reviews and meta-analysis.

| **NO** | **Authors. years** | **Setting** | **Post COVID period-Months** | **Type of case** | **Type of Control** | **Sample Size** | | | **Age** | | **Specimen** | **Quality score** | **Red point score** | **Findings** |
| --- | --- | --- | --- | --- | --- | --- | --- | --- | --- | --- | --- | --- | --- | --- |
|  |  |  |  |  |  | **Cases M/F** | **Control M/F** | **Total M/F** | **Case-Mean (SD)** | **Control- Mean (SD)** |  |  |  |  |
| 1 | (Kovarik, Bileck et al. 2023) | Austria | 7 (3-10) | Post COVID | Healthy Control | 13 4/9 | 13 6/7 | 26 10/16 | 33 (21-53) | 30 (25-43) | Serum | 7.5 | 11.5 | ALB*,Ferritin#,TP# |
| 2 | (Wallis, Heiden et al. 2021) | UK | 3 | Persistent CXR Abnormalities | Complete Resolution | 32 18/14 | 69 36/33 | 101 54/47 | 57.01 (3.26) | 52 (42.5–62.5) | Serum | 7.5 | 11.5 | ALT#,Bilirubin#,Ferritin#,LDH# |
| 3 | (García-Abellán, Fernández et al. 2022) | Spain | 12 | Post COVID | Controls | 14 5/9 | 58 39/19 | 72 44/28 | 60.38 (5.27) | 60 (52-71) | Serum | 4.5 | 12.5 | D-dimer#,Ferritin* |
| 4 | (Agafonova, Elovikova et al. 2024) | Russia | 12 | Post COVID | Controls | 138 27/111 | 87 18/69 | 225 45/180 | 60.77 (4.40) | 59 (44-67) | Blood | 7 | 6 | ALT#,AST#,D-dimer#,LDH*,PLT* |
| 5 | (Alfadda, Rafiullah et al. 2022) | Saudi Arabia | 6 | Post COVID | Controls | NA | NA | 98 NA/NA | 51.34 (18.2) | 46.44 (16.8) | Serum | 3.5 | 13.5 | ALB#,ALP*,ALT*,AST*,Bilirubin*,Bilirubin_1*,D-dimer#,Fibrinogen#,GGT#,LDH*,PLT*,PT#,PTT* |
| 6 | (Al Masoodi, Radhi et al. 2023) | Iraq | NA | Post COVID | Controls | 60 27/33 | 30 8/22 | 90 35/55 | 35.97 (9.17) | 33.23 (6.11) | Serum | 8 | 7.5 | ALB# |
| 7 | (Abdulaziz Alsufyani 2023) | Saudi Arabia | 6 | Post COVID | Controls | 37 37/0 | 35 35/0 | 72 72/0 | 11 (1202) | 10.86 (1089) | Serum | 7 | 6 | ALB#,ALP#,ALT*,AST*,Bilirubin#,GGT*,LDH*,TP* |
| 8 | (Aparisi, Ybarra-Falcón et al. 2021) | Spain | 3 | Post COVID | Controls | 41 11/30 | 29 14/15 | 70 25/45 | 54.9 (10.5) | 54.6 (13.9) | Serum | 4.5 | 11.5 | ALB#,AST#,D-dimer#,Ferritin* |
| 9 | (Aparisi, Ybarra-Falcón et al. 2022) | Spain | 3 | Post COVID | Controls | 10 0/10 | 60 25/35 | 70 25/45 | 46.9 (8.45) | 56.13 (11.9) | Serum | 4.5 | 10.5 | AST*,D-dimer*,Ferritin*,Fibrinogen# |
| 10 | (Belenichev, Kucherenko et al. 2022) | Ukrania | 1 | Post COVID | Controls | NA | NA | 72 NA/NA |  |  | Plasma | 2.5 | 12 | Ferritin#,Ferritin_1# |
| 11 | (Boruga, Septimiu-Radu et al. 2024) | Romania | 6 | Post COVID | Controls | 92 50/42 | 114 59/55 | 206 109/97 | 56.9 (7.6) | 55.2 (8.5) | Serum | 5.5 | 11.5 | ALT#,AST#,D-dimer#,Ferritin* |
| 12 | (Bota, Bratosin et al. 2024) | Romania | 6 | Post COVID | Controls | 117 65/52 | 50 27/23 | 167 92/75 | - | - | Serum | 4.5 | 15.5 | ALB*,ALB_1*,ALP#,ALP_1#,ALT#,ALT_1#,APTT#,APTT_1#,AST#,AST_1#,Bilirubin#,Bilirubin_1#,GGT#,GGT_1#,LDH#,LDH_1#,PT #,PT_1#,TP*,TP_1* |
| 13 | (Cezar, Kundura et al. 2024) | France | 12 | Post COVID | Controls | 19 12/7 | 10 7/3 | 29 19/10 | 68.5 (15.3) | 58.4 (20.8) | Plasma | 3 | 21 | LDH# |
| 14 | (Clemente, Sinatti et al. 2022) | Italy | 3 | Post COVID | Controls | 48 34/14 | 45 22/23 | 93 56/37 | 62.54 (10.18) | 60.62 (17.37) | Serum | 3.5 | 19.5 | ALT#,AST#,Ferritin#,LDH# |
| 15 | (Colarusso, Maglio et al. 2021) | Italy | 3,00 | Post COVID | Healthy Control | 52 32/20 | 17 17/0 | 69 49/20 | 50 (10) | 50 (10) | Plasma | 4.5 | 11.5 | LDH#,LDH_1#,LDH_2#,LDH_3# |
| 16 | (Corrêa, Deus et al. 2022) | Brazil | 11 | Post COVID | Controls | NA | NA | 77 NA/NA | 67.11 (2.99) | 65.31 (4.77) | Plasma | 7.5 | 13.5 | Ferritin# |
| 17 | (Dudar, Loboda et al. 2023) | Ukrania | 8 | HD with Post COVID | HD without Post COVID | 118 60/58 | 82 39/43 | 200 99/101 | 59.5 (52–69) | 47.5 (37.5–60) | NA | 0 | 0 | ALB* |
| 18 | (Dugani, Mehta et al. 2022) | India | NA | Post COVID | Controls | 69 37/32 | 66 45/21 | 135 82/53 | 51.71 (15.9) | 46.33 (18.17) | Serum | 5.5 | 12 | D-dimer#,Ferritin#,LDH# |
| 19 | (Fan, Wong et al. 2022) | Singapore | 12.7+/3.6 | Post COVID | Healthy Control | 39 28/11 | 124 61/63 | 163 89/74 | 43 (32. 56) | 43 (21–65) | Plasma | 5.5 | 10.5 | APTT*,D-dimer#,Factor V*,Factor VIII#,Fibrinogen#,PT# |
| 20 | (Gameil, Marzouk et al. 2021) | Egypt | 6-Mar | Post COVID | Healthy Control | 120 67/53 | 120 69/51 | 240 136/104 | 38.29 (5.27) | 37.25 (4.87) | Serum | 5.5 | 9.5 | ALB*,ALP#,ALT#,AST#,Bilirubin#,D-dimer#,Ferritin#,GGT#,PLT# |
| 21 | (Garcia-Gasalla, Berman-Riu et al. 2023) | Spain | 1.5-3 | Post COVID | Controls | 49 20/29 | 33 22/11 | 82 42/40 | 56.6 (12.5) | 60 (15.5) | Serum | 4.5 | 11.5 | D-dimer#,Ferritin# |
| 22 | (Di Gennaro, Valentini et al. 2022) | Italy | 3 | Post COVID | Controls | 46 22/24 | 29 16/13 | 75 38/37 | 10.5 (6.7) | 10.1 (3) | Plasma | 4.5 | 11.5 | ALB#,ALP#,ALT#,AST#,Bilirubin# |
| 23 | (Guntur, Nemkov et al. 2022) | USA | 1 | Post COVID | Controls | 29 12/17 | 16 8/8 | 45 20/25 | 42 (13) | 60 (14) | Plasma | 0 | 0 | ALB*,ALP#,ALT#,AST#,Bilirubin# |
| 24 | (Gupta, Nicholas et al. 2024) | UK | 7 | PostCOVID | Healthy Control | 21 15/6 | 10 7/3 | 31 22/9 | 54.28 (3.17) | 58 (52–67) | Serum | 4.5 | 13 | ALT#,Bilirubin# |
| 25 | (Ivchenko, Lobzhanidze et al. 2023) | Russia | NA | PostCOVID | Controls | 30 8/22 | 0 0/0 | 60 8/22 | - | - | 0 | 8 | 4 | ALT#,AST#,GGT# |
| 26 | (Kerget, Çelik et al. 2022) | Turkey | 3 | Post COVID | Controls | NA | NA | 0 NA/NA | - | - | 0 | 4.5 | 12.5 | D-dimer#,LDH# |
| 27 | (Kolesova, Vanaga et al. 2021) | Latvia | 3–6 | Post COVID | Controls | 58 31/27 | 17 9/8 | 75 40/35 | 41.2 (13.4) | 42.8 (11.0) | Serum | 5.5 | 8 | ALT#,AST*,Ferritin*,LDH*,PLT* |
| 28 | (Kruger, Vlok et al. 2022) | South Africa | 7 | Post COVID | Healthy Control | 66 21/45 | 29 9/20 | 95 30/65 | 50.85 (4.25) | 52 (41–57) | Serum | 4.5 | 11 | Ferritin*,Ferritin_1* |
| 29 | (Kuchler, Günthner et al. 2023) | Germany | 13.8 | Post COVID with ME/CFS | Controls | 25 6/19 | 16 4/12 | 41 10/31 | 40.6 (12.2) | 44.7 (12.2) | Serum | 5 | 10.5 | D-dimer#,Ferritin# |
| 30 | (Lazebnik, Turkina et al. 2023) | Russia | 4–6 | Post COVID | Controls | 23 13/10 | 60 22/38 | 83 35/48 | 51.71 (12.82) | 51.68 (13.16) | Blood | 4.5 | 22.5 | ALT#,AST# |
| 31 | (Maamar, Artime et al. 2022) | Spain | 3 | Post COVID | Controls | 25 0/25 | 43 0/43 | 68 0/68 | 47.2 (13) | 47.7 (17) | Serum | 5.5 | 15 | D-dimer*,D-dimer_1#,Ferritin*,Ferritin_1#,Fibrinogen#,Fibrinogen_1*,LDH*,LDH_1* |
| 32 | (Magdy, Eid et al. 2022) | Egypt | NA | Post COVID | Controls | 45 15/30 | 45 16/29 | 90 31/59 | 43 (15.85) | 45.71 (13.89) | Serum | 4.5 | 12.5 | Ferritin# |
| 33 | (Martone, Tosato et al. 2022) | Italy | 3 | Post COVID sarcopenia | Controls | 106 42/64 | 435 225/210 | 541 267/274 | 59.9 (15.9) | 51.5 (14.6) | Serum | 5.5 | 11 | ALB* |
| 34 | (Meisinger, Goßlau et al. 2022) | Germany | 9.3 | Post COVID | Controls | 72 0/72 | 80 0/80 | 152 0/152 | 45.41 (16.26) | 45.11 (19.25) | blood | 8.5 | 11 | PLT*,PLT_1* |
| 35 | (Nádasdi, Sinkovits et al. 2022) | Hungary | NA | Post COVID | Controls | 26 15/11 | 39 0/39 | 65 15/50 | 44.74 (5.04) | 37.0  (34–40) | Serum | 4.5 | 11.5 | ALB#,ALB_1# |
| 36 | (Paniskaki, Goretzki et al. 2023) | Germany | 5 | Post COVID | Controls | 17 8/9 | 13 11/2 | 30 19/11 | 11 (-) | 12 (-) | Blood | 4.5 | 11.5 | ALT*,AST*,D-dimer*,GGT* |
| 37 | (Fernandez-de-las-Peñas, Notarte et al. 2024) | Spain | 6 | Post COVID | Controls | 300 140/160 | 112 73/39 | 412 213/199 | 63 (15) | 59.5 (17) | Serum | 5.5 | 10.5 | ALT*,ALT_1*,ALT_2*,ALT_3*,AST*,AST_1#,AST_2*,AST_3*,D-dimer#,D-dimer_1*,D-dimer_2*,D-dimer_3*,LDH*,LDH_1*,LDH_2*,LDH_3*,PLT*,PLT_1*,PLT_2*,PLT_3* |
| 38 | (Portacci, Amendolara et al. 2024) | Italy | 4.3 | Post COVID | Controls | 318 170/148 | 119 78/41 | 437 248/189 | 58 (51–66) | 58 (49–67) | Serum | 4.5 | 14 | LDH#,PLT# |
| 39 | (Poyatos, Luque et al. 2024) | Spain | 3 | Post COVID | Healthy Control | 29 24/5 | 31 20/11 | 60 44/16 | 64.6 (13.8) | 58.6 (8.21) | Plasma | 5 | 14 | Ferritin#,Ferritin_1#,Ferritin_2#,Fibrinogen#,Fibrinogen_1*,Fibrinogen_2#,LDH#,LDH_1#,LDH_2# |
| 40 | (Radzina, Putrins et al. 2022) | Latvia | 6.4 | Post COVID | Controls | 56 28/28 | 34 13/21 | 90 41/49 | 41.6 (13.4) | 39.5 (12.9) | Plasma | 4.5 | 7 | ALT#,AST#,GGT*,LDH* |
| 41 | (Díaz-Salazar, Navas et al. 2022) | Spain | 3 | Post COVID | Controls | 36 11/25 | 85 42/43 | 121 53/68 | 46.7 (14) | 45.1 (17) | Serum | 7.5 | 12 | ALB#,ALB_1*,ALB_2#,D-dimer*,D-dimer_2#,Ferritin*,Ferritin_2#,Fibrinogen#,Fibrinogen_1*,Fibrinogen_2#,LDH#,LDH_1*,LDH_2* |
| 42 | (Sasso, Muraki et al. 2022) | Australia | 6.7 | Post COVID | Controls | 5 2/3 | 5 1/4 | 10 3/7 | 50.8 (8.76) | 39.8 (14.77) | Blood | 6.5 | 8.5 | PLT* |
| 43 | (Sibila, Perea et al. 2022) | Spain | 6 | Post COVID DLCO (< 80% ref,) | Post COVID DLCO (≥ 80% ref,) | 90 59/31 | 125 71/54 | 215 130/85 | 56.9 (12.7) | 65.3 (10.4) | Plasma | 8.5 | 11.5 | D-dimer*,Ferritin#,LDH*,PLT* |
| 44 | (Sommen, Havdal et al. 2023) | Norway | 7 (6-12) | Post COVID | Controls | 367 145/222 | 81 30/51 | 448 175/273 | 18 (6) | 18 (5) | Plasma | 7.5 | 14.5 | ALT#,Ferritin#,PLT#,PLT_1# |
| 45 | (Stavileci, Özdemir et al. 2022) | Turkey | 6 | Post COVID fragmented QRS | Non-de-novo fragmented QRS | 102 44/58 | 146 50/96 | 248 94/154 | 36.47  (8.72) | 33.64  (9.22) | Serum | 5.5 | 12.5 | ALT#,AST*,D-dimer#,Ferritin#,LDH# |
| 46 | (Stufano, Isgrò et al. 2023) | Italy | 10 | Post COVID | Controls | 34 15/19 | 47 33/14 | 81 48/33 | 50.9 (9.4) | 53.8 (12.6) | blood | 5 | 15 | PLT* |
| 47 | (Sumbalova, Kucharska et al. 2022) | Slovakia | NA | Post COVID | Healthy Control | 14 8/6 | 15 6/9 | 29 14/15 | 51.3 (2.3) | 51.3 (2.3) | blood | 4 | 14 | PLT*,PLT_1* |
| 48 | (Sumbalová, Kucharská et al. 2022) | Slovakia | 2 | Post COVID | Healthy Control | 10 3/7 | 15 6/9 | 25 9/16 | 59.9 (5.4) | 51.3 (2.3) | Plasma | 4.5 | 14 | ALB*,GGT#,TP* |
| 49 | (Taha, Samaan et al. 2021) | Egypt | 6 | Post COVID arthritis | No arthritis | 37 23/14 | 63 38/25 | 100 61/39 | 63.06 (12.33) | 50.93 (19.95) | Serum | 4.5 | 12 | ALB*,ALT#,AST#,D-dimer#,Ferritin#,LDH#,PLT# |
| 50 | (Teng, Song et al. 2023) | China | 6 | Post COVID With kidney involvement | Without kidney involvement | 8 3/5 | 39 27/12 | 47 30/17 | 42 (15.18) | 49.1 (14.36) | Serum | 5 | 10.5 | ALB#,ALT#,AST*,Ferritin#,GGT#,PLT# |
| 51 | (Torki, Hoseininasab et al. 2024) | Iran | At least 3 | Post COVID | Controls | 88 43/45 | 96 55/41 | 184 98/86 | 47.45 (3.31) | 43 (37.25–58) | Blood | 4.5 | 9 | ALT*,AST#,Ferritin*,LDH#,PLT#,PTT# |
| 52 | (Fogarty, Townsend et al. 2021) | Ireland | 2 | fatigued | non-fatigued | 67 22/45 | 61 37/24 | 128 59/69 | 49.3 (14.3) | 49.7 (16) | Serum | 5.5 | 10.5 | LDH* |
| 53 | (Wu, Deng et al. 2021) | China | 6 | Post COVID with cardiac injury | Post COVID without cardiac injury | 24 14/10 | 26 7/19 | 50 21/29 | 59.63  (10.43) | 48.73  (9.07) | Serum | 7.5 | 12.5 | ALT#,AST#,D-dimer#,LDH# |
| 54 | (Parás-Bravo, Fernández-de-las-Peñas et al. 2024) | Spain | 12 (8) | Post COVID fatigue | No Fatigue | NA | NA | 76 NA/NA | 69.0 (10.5) | - | 0 | 6.5 | 10 | ALT*,ALT_1*,ALT_2*,ALT_3*,AST*,AST_1#,AST_2*,AST_3*,D-dimer#,D-dimer_1*,D-dimer_2*,D-dimer_3*,LDH*,LDH_1*,LDH_2*,LDH_3*,PLT*,PLT_1*,PLT_2*,PLT_3* |
| 55 | (Frontera, Betensky et al. 2024) | USA | 18 | Post COVID cognitive symptoms | without cognitive symptoms | 76 33/43 | 203 24/179 | 279 57/222 | 57 (47–64) | 59 (52–69) | Serum | 5.5 | 13 | D-dimer#,D-dimer_1#,Ferritin#,Ferritin_1#,Fibrinogen#,Fibrinogen_1#,LDH#,LDH_1# |
| 56 | (Kalinskaya, Vorobyeva et al. 2023) | Russia | NA | Post COVID | Controls | 31 8/23 | 27 10/17 | 58 18/40 | 49 (47-55.5) | 48 (43.5-54.5) | Blood | 4.5 | 17 | ALT#,aPTT#,AST#,D-dimer#,PLT*,PT* |
| 57 | (Kankaya, Yavuz et al. 2023) | Turkey | 5 | Post COVID | Healthy Controls | 53 21/32 | 30 18/12 | 83 39/44 | 37 (-) | 40 (-) | Blood | 3 | 18 | D-dimer*,Ferritin*,Fibrinogen# |
| 58 | (Paris, Palomba et al. 2023) | Italy | 2 | Post COVID | Healthy Controls | 38 35/3 | 38 35/3 | 76 70/6 | 58.82 (10.08) | 57.93 (11.23) | Serum | 8 | 11 | ALB#,ALT#,AST#,D-dimer#,PLT* |
| 59 | (Petramala, Sarlo et al. 2023) | Italy | At least 2 | Post COVID with pulmonary embolism | Healthy controls | 18 6/12 | 44 21/23 | 62 27/35 | 64.3 (5.3) | 62 (7.2) | Serum | 5 | 14 | aPTT#,D-dimer#,Fibrinogen#,PT# |
| 60 | (Vollrath, Matits et al. 2023) | Germany | 3.8 ± 2.68 | Post COVID | Controls | 59 30/29 | 31 12/19 | 90 42/48 | 34.5 (12.2) | 31.9 (10.4) | Blood | 6 | 16 | ALT#,PT#,TP* |
| 61 | (Yamamoto, Otsuka et al. 2023) | Japan | At least 6 | Post COVID with ME/CSF | No fatigue | 50 24/26 | 95 38/57 | 145 62/83 | 42 (30.3–51.8) | 43 (29.5-51) | Blood | 7.5 | 12 | ALB#,ALP*,ALT#,AST#,Bilirubin#,Ferritin#,Fibrinogen*,PLT#,TP# |

*: Indicates that patients have reduced level of the measured metabolite compared to healthy control (negative SMD)

^#:^ Indicates that patients have increased level of the measured metabolites compared to healthy control (positive SMD)

TRP: Tryptophan. KYN: Kynurenine. KA: Kynurenic acid. 3HK: 3-Hydroxykynurenine. AA: Anthranilic acid. 3HA: 3-Hydroxyanthranilic acid. XA: Xanthurenic acid. QA: Quinolinic acid. PA: Picolinic acid. HPLC: High performance liquid chromatography. HPLC-MS/MS: High performance liquid chromatography with tandem mass spectrometry. LC-MS/MS: Liquid chromatography with tandem mass spectrometry. UPLC-MS/MS: Ultra performance liquid chromatography with tandem mass spectrometry. LC: Liquid chromatography. HPLC-UV: High perfomance liquid chromatography- Ultra-violate

**ESF. Table 6**. Results of Meta-regression

| Variables | No. of Studies | Covariates | 1-sided p-value | Z-Value |
| --- | --- | --- | --- | --- |
| AST/ALT ratio | 5 | Patients on Immunomodulatory Drugs | 2.10 | 0.017 |
| ALT | 5 | Smoking | 0.023 | 1.99 |
| LDH | 8 | Hospitalized Patients During Acute COVID | 0.0006 | -3.24 |
|  | 21 | Age | 0.015 | 2.16 |
|  | 8 | BMI | 0.007 | 2.46 |
|  | 10 | Smoking | 0.035 | 1.80 |
|  | 23 | Quality Control | 0.027 | -1.92 |
|  |  | Red Points | 0.006 | 2.46 |
| Ferritin | 26 | Recruited Under 3 Months Post-Acute COVID | 0.037 | 1.78 |
| Platelet count | 6 | Hospitalized Patients During Acute COVID | 0.036 | -1.79 |
|  | 10 | Age | 0.045 | -1.70 |
|  | 16 | Recruited Under 3 Months Post-Acute COVID | 0.045 | -1.69 |
| Albumin | 5 | Patients on Immunomodulatory Drugs | 0.0049 | 2.58 |
|  |  | Smoking | 0.049 | 1.65 |
| Prothrombin time | 5 | Sample size | 0.030 | 1.88 |
| Total Protein | 5 | Patients on Immunomodulatory Drugs | 0.003 | 2.75 |
|  |  | Smoking | 0.049 | 1.65 |
| D-Dimer | 26 | Latitude | 0.012 | -2.25 |
|  | 11 | Hospitalized Patients During Acute COVID | 0.039 | -1.76 |
|  | 8 | Smoking | 0.042 | 1.72 |
| Fibrinogen | 11 | Recruited Over 6 Months Post-Acute COVID | 0.035 | 1.80 |
|  | 10 | Sample size | 0.012 | 2.25 |
|  | 7 | BMI | 0.000 | 4.04 |

ALT: Alanine aminotransferase, Aspartate transaminase, ALP: Alkaline phosphatases, GGT: Gamma-glutamyl transferase, LDH: Lactate dehydrogenase.

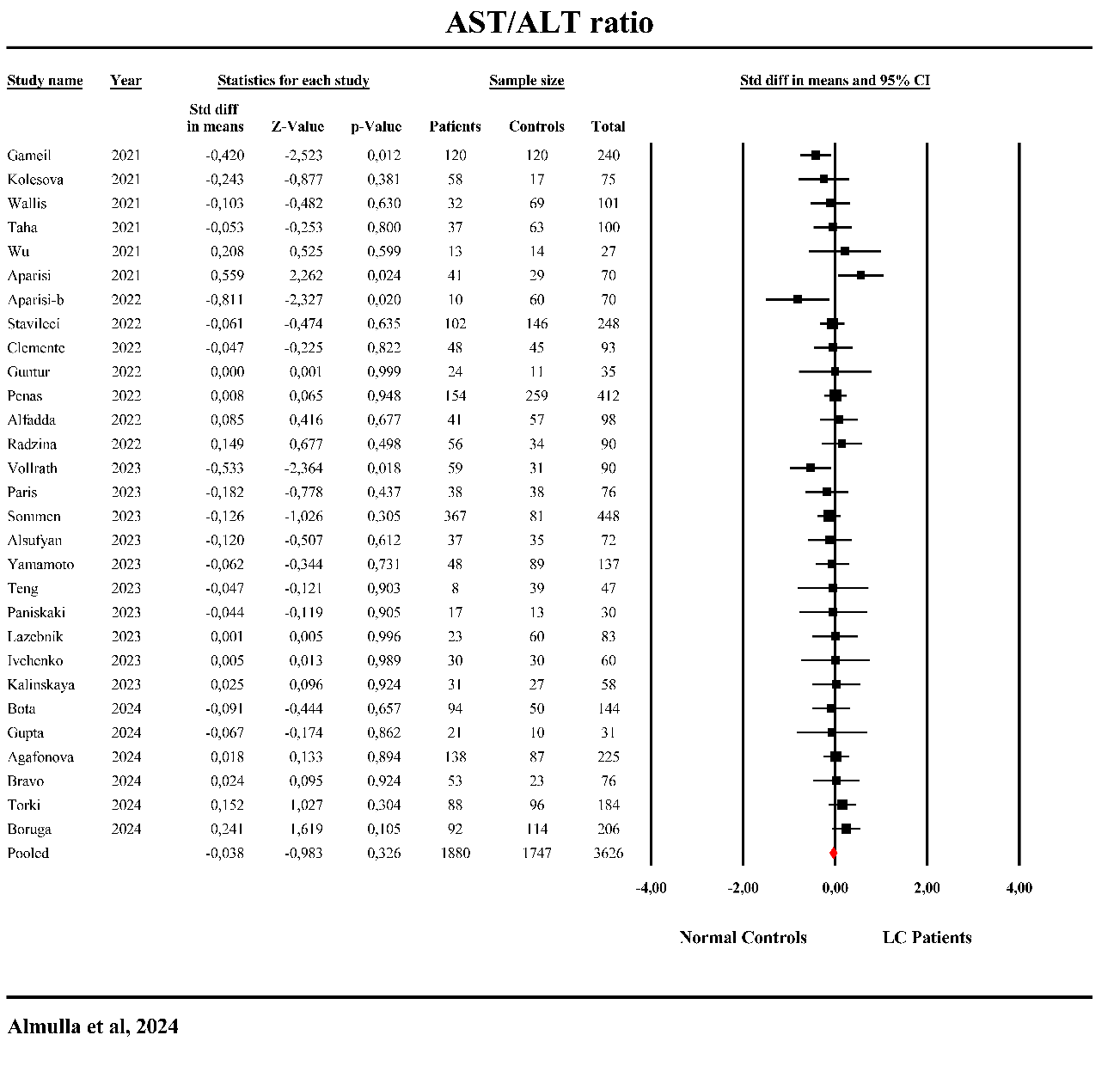

**ESF. Figure 1**. Forest plot of Aspartate aminotransferase (AST)/Alanine aminotransferase (ALT) in the patients with Long COVID (LC) versus normal controls.

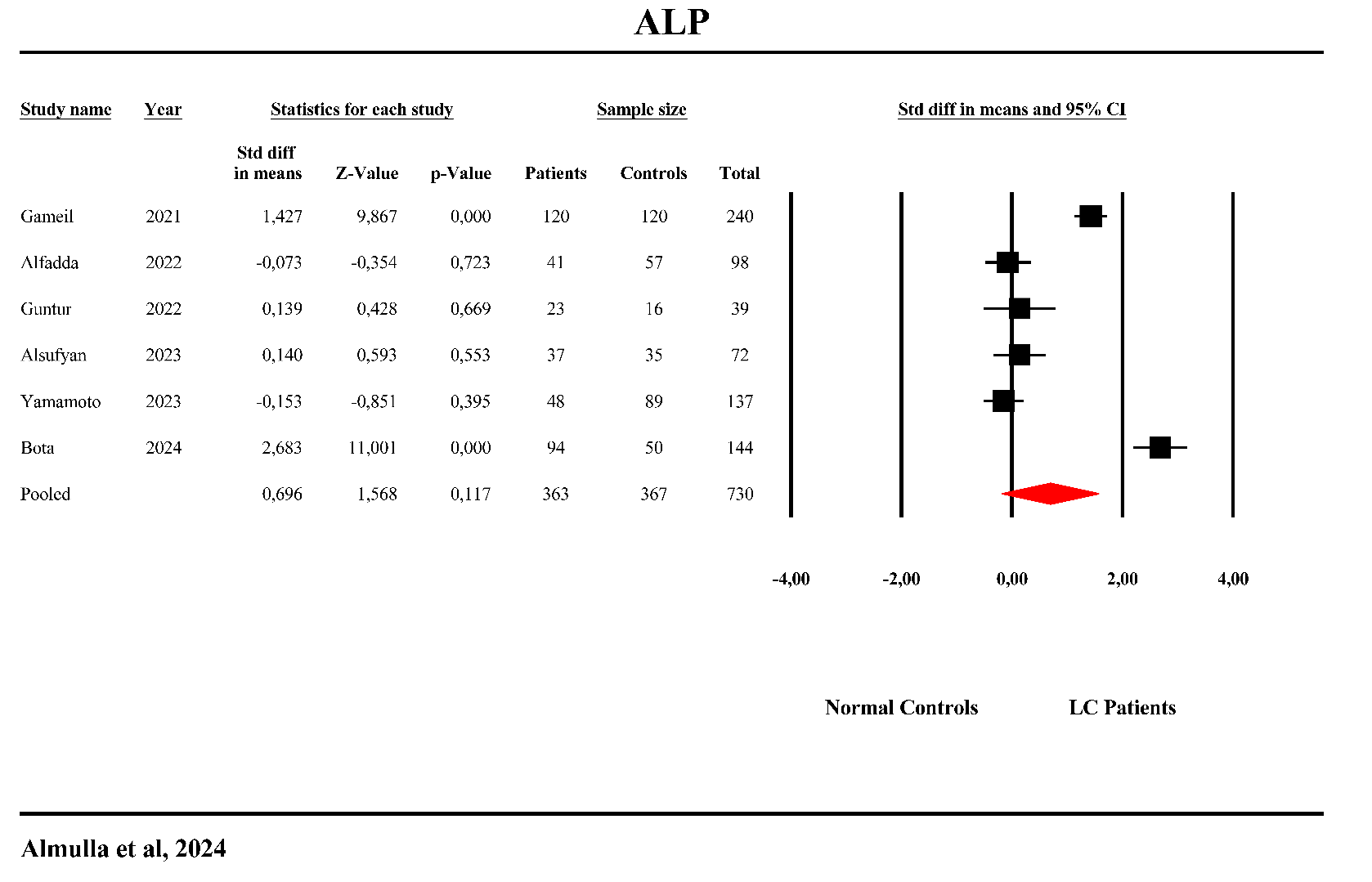

**ESF. Figure 2**. Forest plot of Alkaline Phosphatase (ALP) in the patients with Long COVID (LC) versus normal controls.

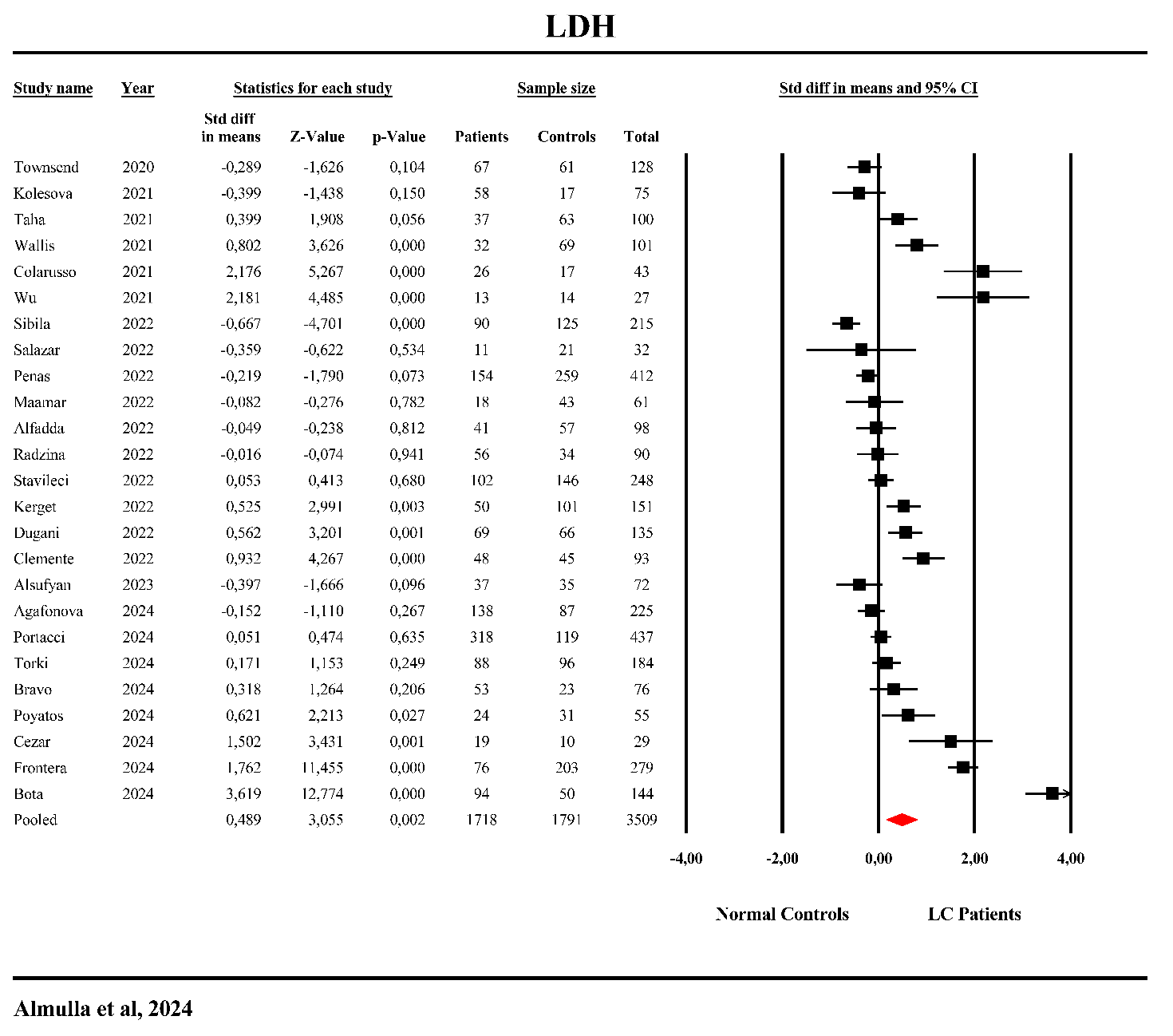

**ESF. Figure 3**. Forest plot of Lactate dehydrogenase (LDH) in the patients with Long COVID (LC) versus normal controls.

.

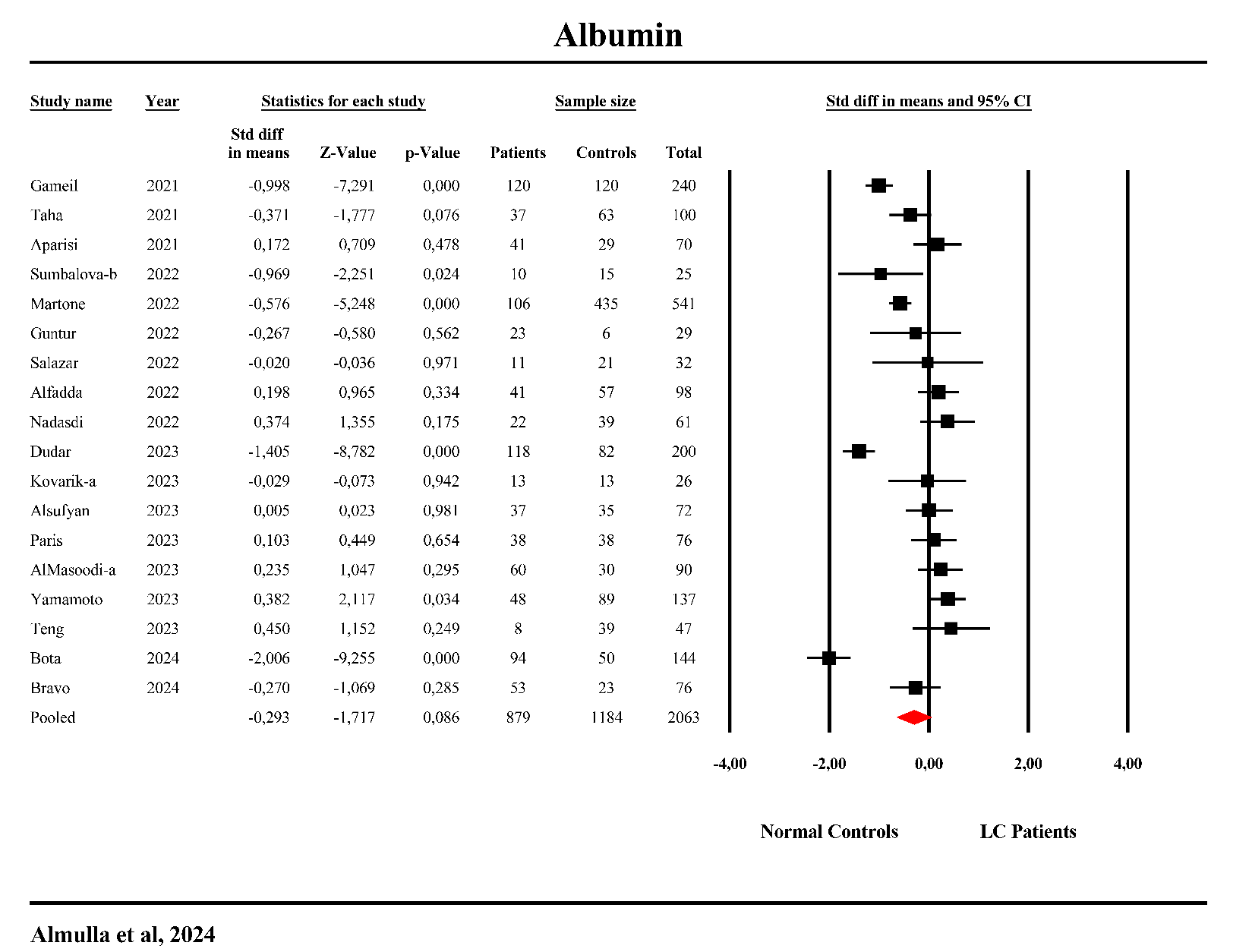

**ESF. Figure 4**.Forest plot of albumin in patients with Long COVID (LC) versus normal control.

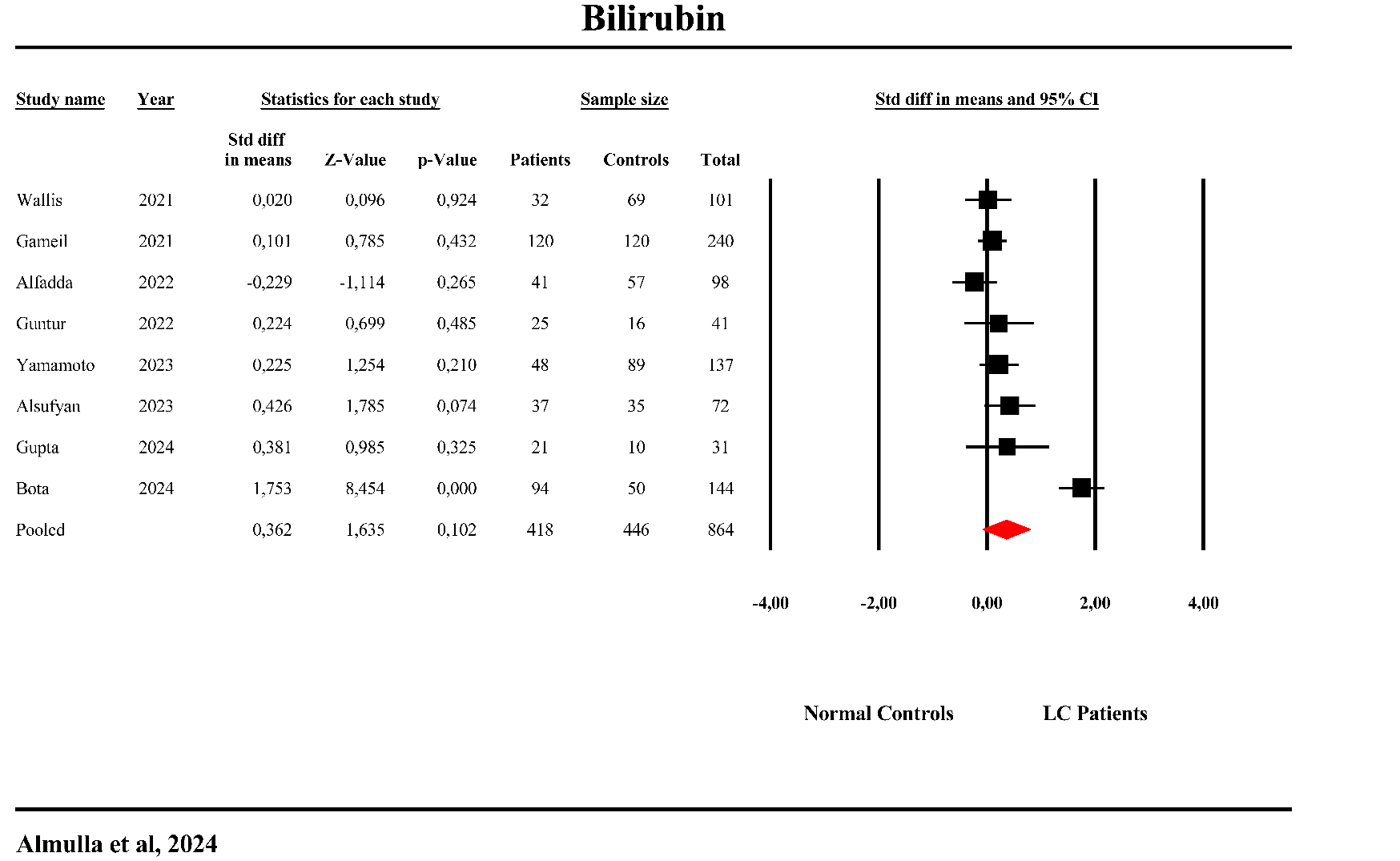

**ESF. Figure 5**. Forest plot of bilirubin in patients with Long COVID (LC) versus normal control.

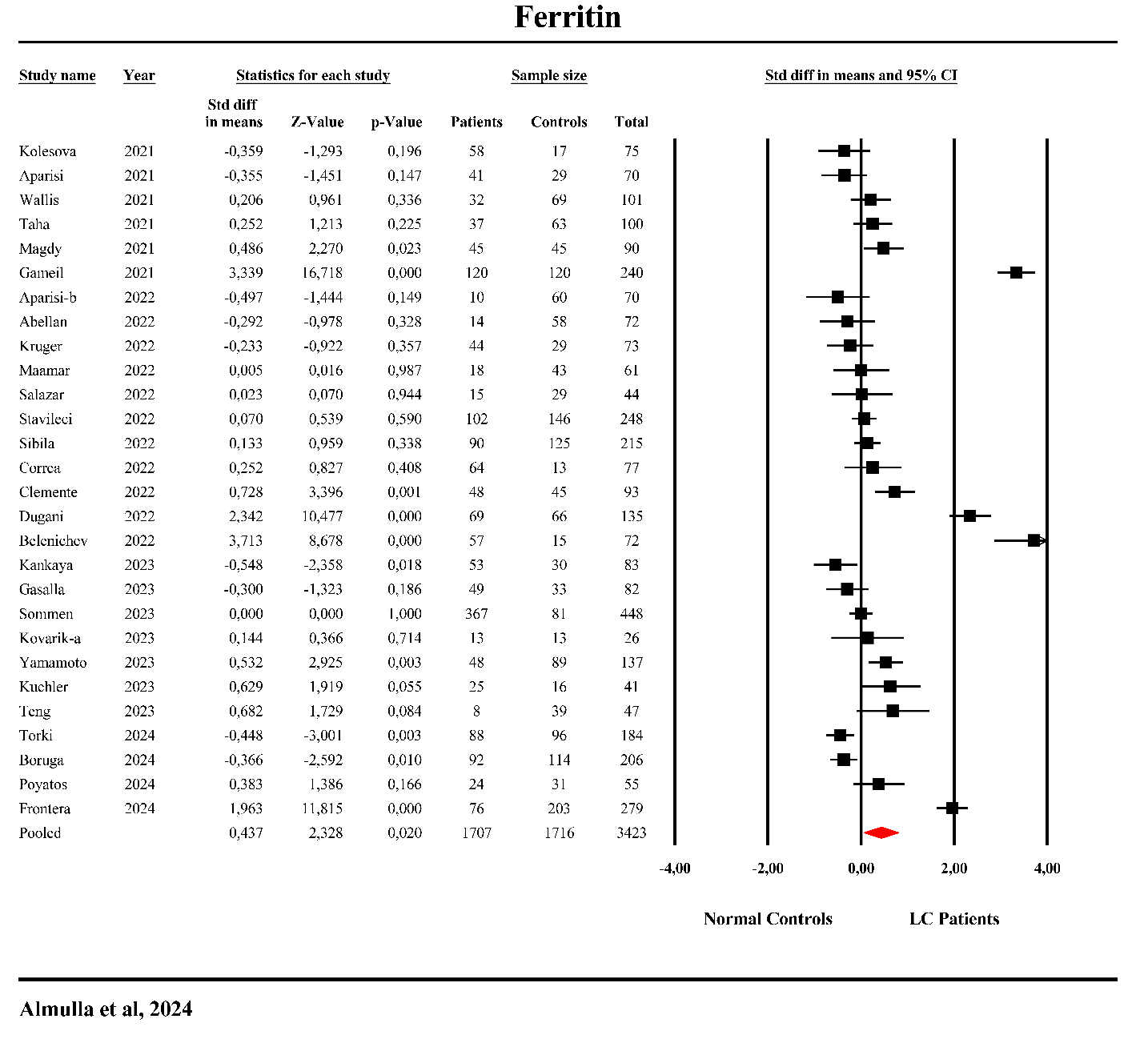

**ESF. Figure 6**. Forest plot of ferritin in patients with Long COVID (LC) versus normal control.

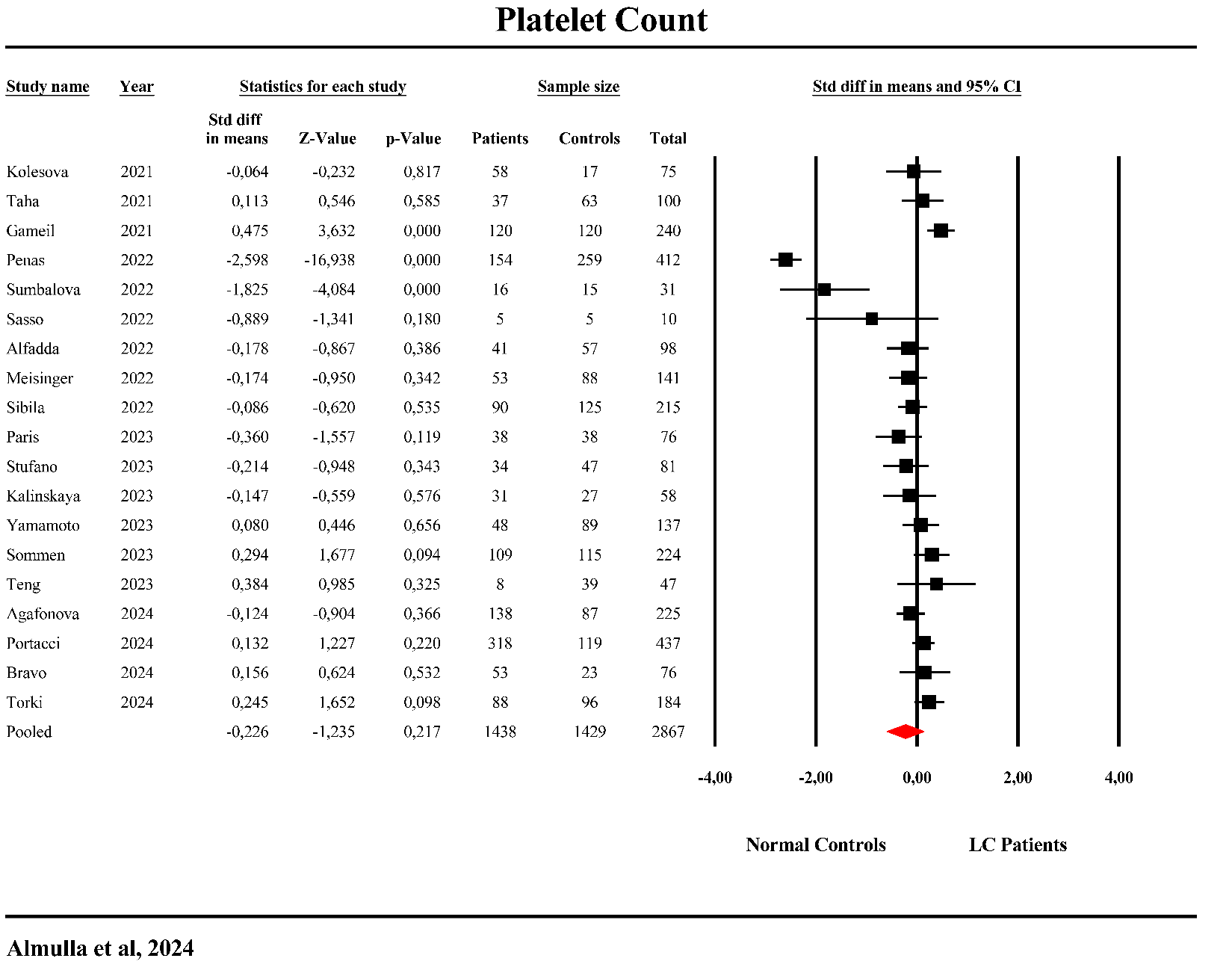

**ESF. Figure 7**. Forest plot of platelet count in patients with Long COVID (LC) versus normal control.

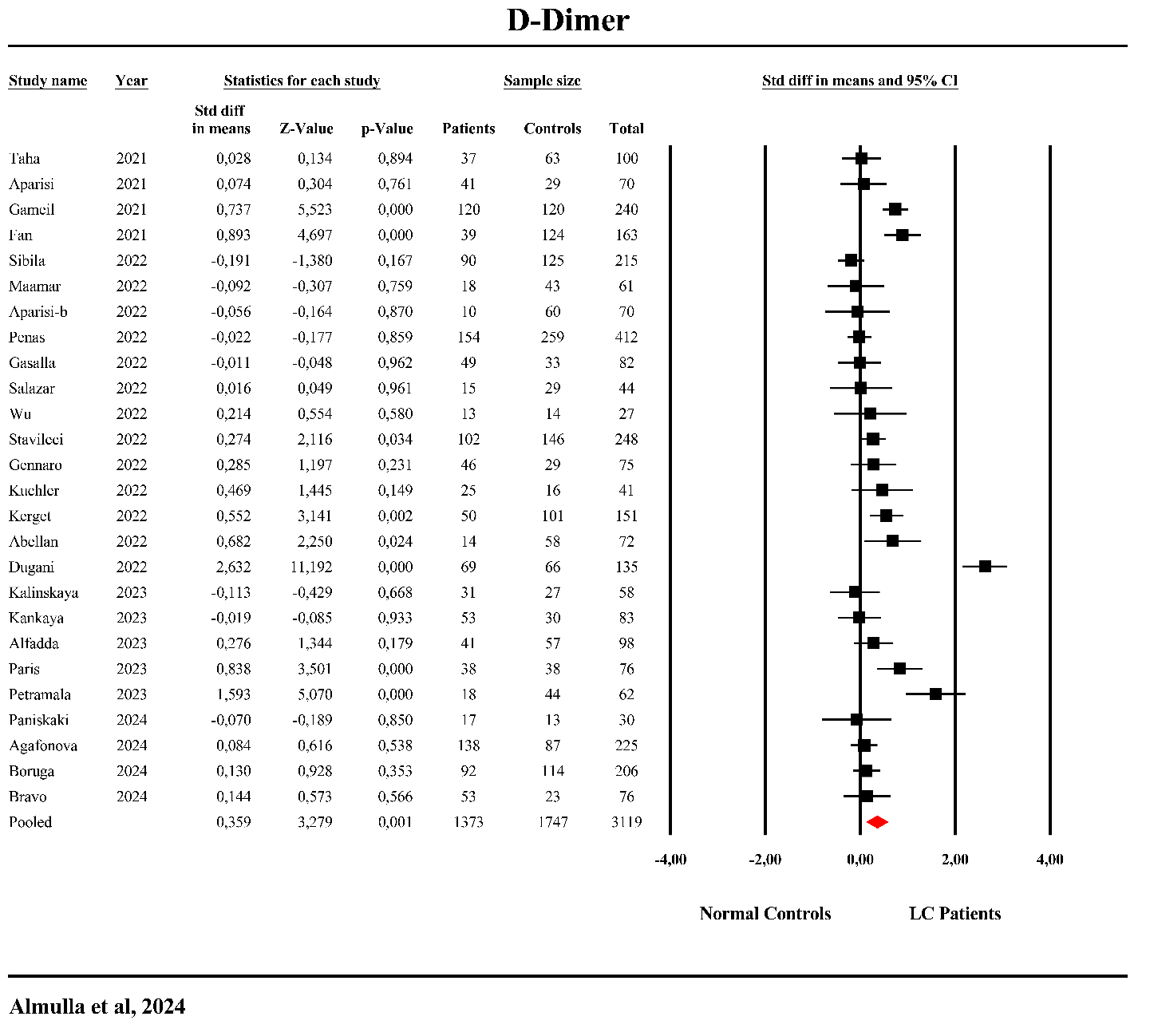

**ESF. Figure 7**. Forest plot of D-dimer in patients with Long COVID (LC) versus normal control.

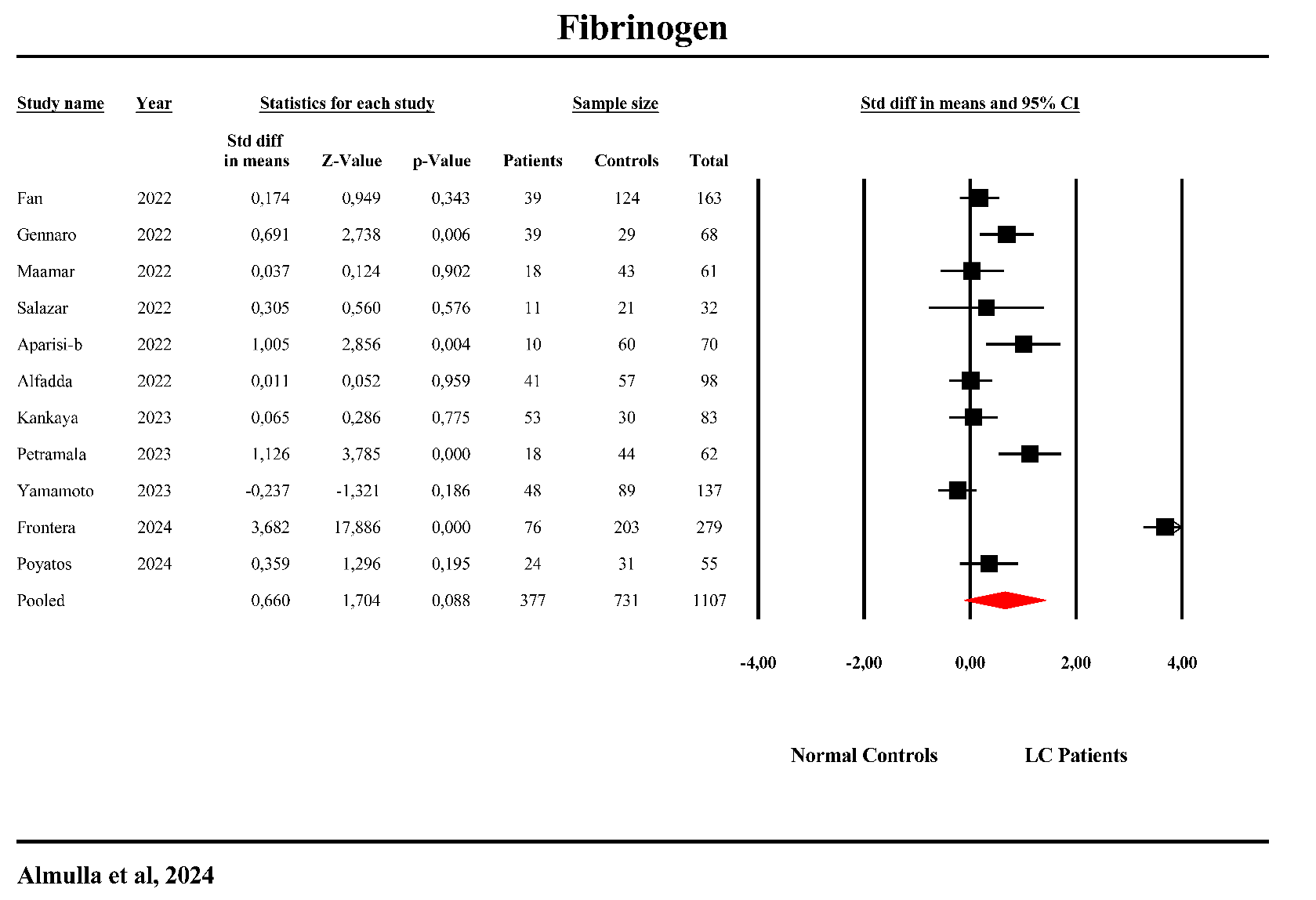

**ESF. Figure 8**. Forest plot of fibrinogen in patients with Long COVID (LC) versus normal control.
